# Measuring adult mortality from mobile phone surveys in Burkina Faso, Malawi and the Democratic Republic of the Congo

**DOI:** 10.1101/2025.02.07.25321855

**Authors:** Kassoum Dianou, Bruno Masquelier, Shammi Luhar, Bruno Lankoandé, Ashira Menashe Oren, Abdramane Soura, Hervé Bassinga, Malebogo Tlhajoane, Boniface Dulani, Pierre Akilimali, Georges Reniers

## Abstract

In many low and middle-income countries, adult mortality estimates are derived from surveys and censuses conducted through face-to-face interviews. These interviews can be time-intensive and are often impractical during health crises or humanitarian emergencies. The expansion in cellphone ownership and network coverage has created new opportunities for collecting demographic data through mobile phone surveys, but our understanding of selection biases and reporting errors of such data remains incomplete. This study reports on adult mortality estimates obtained through mobile phone surveys conducted in Burkina Faso, Malawi, and the Democratic Republic of the Congo in 2021 and 2022. To mitigate respondent fatigue and network interruptions, we used a shortened version of the set of questions generally used in surveys to ask about the survival of respondents’ siblings. We found substantial differences between mortality estimates obtained from mobile phone interviews and those from face-to-face demographic surveys. Mortality rates from the mobile phone surveys were also approximately half those expected from United Nations estimates. We attribute this underestimation to inaccuracies in reporting of ages and the timing of sibling deaths in the shortened sibling instrument. After imputing ages and dates based on full sibling histories collected in previous face-to-face surveys, mortality rates were more consistent with other data sources. Mobile phone surveys are promising for the measurement of adult mortality in settings where face-to-face surveys are hindered, but they are susceptible to reporting errors. More research is needed on the best set of questions to use for capturing recent adult deaths.

## 1. Introduction

Comprehensive systems of death registration are the ideal source to obtain accurate and timely information on mortality and causes of death, but in many countries these systems remain deficient [1,2]. The completeness of death registration is particularly low in Sub-Saharan Africa, with the exception of a few countries, such as Zimbabwe and South Africa [3]. In the absence of efficient systems of death registration, surveys and censuses remain the primary sources of mortality data. By pooling data sources together, it is now possible to monitor trends in child mortality with a relatively good precision [4]. Data is scarcer for adults. In particular, retrospective reports on the survival of siblings or parents collected in Demographic and Health Surveys [DHS] are one of the most widely used data source on adult mortality [5], alongside reports on recent household deaths collected in censuses. While these sources facilitate the reconstruction of mortality trends, data collection is usually organized through face-to-face interviews, which limits their potential for capturing rapid changes in mortality, as nationwide surveys or censuses are carried out irregularly. These large operations are also challenging during humanitarian and public health emergencies. For instance, during the Ebola and COVID-19 health crises, many survey programs and census operations had to be temporarily suspended [6].

Alternative approaches to data collection are needed to monitor mortality more frequently and in resource-constrained environments where in-person surveys are restricted. The expansion of mobile phone ownership and network coverage opens the door to mobile phone data collection in such contexts. Telephone interviews can be more cost-effective, offer greater flexibility in questionnaire design, quicker deployment, and consequently make it possible to collect data more frequently than traditional face-to-face surveys [7–11]. Until recently, nationwide mobile phone surveys (MPS) were uncommon in low- and middle-income countries, but the COVID-19 pandemic greatly accelerated the adoption of [12–16]. So far, few MPS were designed to track changes in mortality. In Monrovia (Liberia), a study conducted by *Médecins Sans Frontières* showed that mobile phone data collection was a viable alternative for measuring excess mortality related to the Ebola crisis [17]. Other MPS conducted in India have successfully documented excess mortality during the COVID-19 pandemic [18,19]. These surveys were based on reports of household deaths, however, and experience is lacking around the feasibility of using other types of survey instruments in MPS, including sibling survival histories (SSH). In full SSH, respondents are asked about the survival status of each of their maternal siblings, their current age if alive and age at the time of death, as well as the time elapsed since death if the sibling is deceased [20]. Although SSH are a well-established mortality measurement tool, this approach had never been tested within a MPS prior to this study.

Collecting sibling survival data over the phone presents a number of challenges. First, it may be desirable to keep questionnaires short, while collecting full SSH can take time in settings where fertility is high. Revisions to the standard SSH module might thus be required. Second, MPS could lead to more misunderstandings between interviewers and respondents, or lower commitment to the survey. A recent study in six sub-Saharan African countries showed that age misreporting errors were more frequent in MPS than in recent household surveys and censuses [21]. Third, respondents in telephone surveys may exhibit some reservation towards certain questions [22], and be more hesitant to provide sensitive information about the deaths of their relatives over the phone. Encouragingly, a randomized trial in Malawi suggested that respondents in a telephone survey were as cooperative when asked about the mortality of relatives as when asked about their recent economic activity, and that questions about the death of close relatives rarely generated negative reactions [23]. Previous research has also demonstrated that MPS can be better suited for addressing sensitive issues, in part because respondents are free to choose where they wish to take the call [9,22,24]. Fourth, the representativeness of MPS respondents could be compromised, especially in places where mobile phone coverage is not universal [7,25,26]. Respondents in MPS are more likely to be young, well-educated, urban, and affluent, in part due to limited telephone coverage and lower purchasing power among poor populations [27]. Stratified quota sampling and post-stratification weighting can help mitigate selection bias, but mortality rates may remain inaccurate if there is a causal link between mobile phone ownership and the survival of close relatives through improved care access and information.

This study reports on siblings survival history estimates of adult mortality from three MPS conducted in Burkina Faso, Malawi, and the Democratic Republic of Congo (DRC). We used a shortened version of the standard sibling survival history used in the DHS. More specifically, we asked respondents to report on the total number of their maternal siblings, the number of such siblings who were still alive, and those who had died, with additional questions for those who had died since the beginning of 2019. Here, we first present an approach to imputing sibling histories collected with this shortened list of questions. We then evaluate the quality of the summary sibling data and compare adult mortality estimates obtained from the MPS to those from the World Population Prospects (WPP), as well as previous DHS and census data.

## 2. Data and methods

### 2.1 Data collection

The Rapid Mortaliy Mobile Phone Surveys Project (RaMMPS, https://www.lshtm.ac.uk/rammps/) is a multi-country survey program to develop and apply methods for measuring mortality via MPS. In this study, we use data from the DRC, Malawi and Burkina Faso that included a SSH module. In Burkina Faso, the survey was coordinated by the *Institut Supérieur des Sciences de la Population* (ISSP) of Joseph Ki-Zerbo University; in DRC by the *Ecole de Santé Publique de l’Université de Kinshasa* (UNIKIN); and in Malawi, it was a joint collaboration between the *Malawi Epidemiology and Interventions Research Unit* (MEIRU) and the *Institute for Public Opinion Research* (IPOR).

Interviews were conducted via mobile phones, with responses recorded on the SurveyCTO platform. Following a few eligibility screening questions, participants provided verbal informed consent. The study protocol received approval from the national health ethics committee for Burkina Faso, and the LSHTM Research Ethics Committees (protocol #26393) for Malawi and DRC. Furthermore, the protocols for Malawi and the DRC have been approved by the University of Malawi Research Ethics Committee (reference: P.07/21/76) and the Ethics Committee of the School of Public Health, University of Kinshasa (reference: ESP/CE/112/2021), respectively. The characteristics of the three MPS are summarized in Table 1.

**Table 1:** Characteristics of mobile phone surveys in the three countries covered.

|  | Burkina Faso | DRC | Malawi |
| --- | --- | --- | --- |
| Data collection period | 09/2021 – 10/2022 | 08/2021 – 08/2022 | 01/2022 – 07/2023 |
| Sample size | 21 339 | 11 924 | 14 663 |
| Coverage | National | Kinshasa and North Kivu provinces | National |
| Recruitment strategy | Numbers based on a previous face-to-face survey [EHCVM] and random digit dialing | Random digit dialing | Random digit dialing |
| Target population for post-stratification | Population of heads of households [for the EHCVM arm] and population aged 15-64 (for random digit dialing) in the 2019 census | Population aged 18 and above in the MICS 2017-2018 | Population aged 18 and above in the DHS 2015-2016 |
| Study team | ISSP, Louvain University and LSHTM | UNIKIN and LSHTM | MEIRU, IPOR and LSHTM |

Notes: DHS: Demographic and Health Survey. EHCVM: Enquête harmonisée sur les conditions de vie des ménages. IPOR: Institute for Public Opinion Research. ISSP: Institut Supérieur des Sciences de la Population of Joseph Ki-Zerbo University. LHSTM: London School of Hygiene & Tropical Medicine MEIRU: Malawi Epidemiology and Interventions Research Unit. MICS: Multiple Indicator Cluster Survey. UNIKIN: Université de Kinshasa.

In the three MPS, respondents were informed that they would receive call credit upon completing the interview to improve survey participation [28–30]. Interviewers were instructed to make repeated attempts to contact individuals over a period of at least seven days at various times during the day before determining that respondents were unreachable.

Across the three countries, we opted for different sampling designs and introduced variations in the questionnaires to test for respondent selectivity based on the recruitment approach [Table 1]. In Burkina Faso, data collection was carried out through quarterly cross-sectional surveys conducted over a 12-months period, to mitigate biases associated with seasonality of mortality. Two distinct sampling strategies were employed. The first sub-sample, referred to as the EHCVM arm, comprised approximately 6,000 individuals selected based on telephone numbers collected during a prior face-to-face survey known as the Harmonized Household Living Conditions Survey (EHCVM for *Enquête Harmonisée sur les Conditions de Vie des Ménages*). For the MPS, we re-contacted each head of household who had been previously interviewed in the face-to-face EHCVM survey and provided a mobile phone number. Additionally, with the consent of the head of the household, a woman of reproductive age (15–49 years) residing in the household was randomly selected for inclusion. The second sub-sample, referred to as the RDD arm, included approximately 9,000 individuals randomly selected by drawing telephone numbers through random digit dialing (RDD). We set quotas for a number of population strata to ensure the representativeness on age, gender, and place of residence, using nationwide distributions based on the latest national census data [31]. Numbers were randomly generated accounting for the prefixes used by the major cell phone operators in the country (MoovAfrica, Telecel Faso, and Orange). Numbers were screened by a company dialing each number to verify their functionality. More details on the Burkina Faso MPS are provided elsewhere [32].

In DRC and Malawi, we only used RDD. In these two countries, we worked with a third party to randomly draw numbers, based on the mobile phone structure used by the major operators in these countries. In the DRC, these were further restricted to numbers that were active in the Kinshasa and North Kivu in the recent past. As in Burkina Faso, we also resorted to quotas informed by national or provincial distribution of the population on a number of key attributes.

To contain the length of the interviews, we developed a shortened version of the full SSH module used in DHS. The questions focused on the total number of siblings, the count of siblings currently alive, and those who had passed away since the start of 2019. We collected supplementary details only for recent deaths, including the date, circumstances, and location of the death (see Table S3 in appendix). The questionnaires for Malawi and DRC were less developed than in Burkina Faso on this module; no questions were asked about ages at death in DRC, and the data do not allow disaggregation between brothers and sisters in Malawi (Table S4). In what follows, we therefore present results by sex for the first two countries, and results for both sexes combined in Malawi.

### 2.2 Estimating adult mortality from MPS

#### 2.2.1 Post-stratification weighting

As indicated earlier, mobile phone ownership is generally higher among individuals with higher socioeconomic status and education [9,32]. In addition, households with more members are more likely to be included in samples of surveys conducted over the phone because there is a higher chance that someone can pick up the call, or because there are more phones available in larger households [34]. To correct for these two sources of errors, we used post-stratification weighting [7]. Weights were calculated using an iterative proportional fitting (a.k.a. raking) procedure to ensure that the marginal distribution of the weighting variables matched the distribution in a target population, such as the latest DHS or census (see Table 1). The selection of target populations in our weighting process varied slightly by country due to the availability of variables and of different external sources (see Table S5 in appendix). In DRC, for example, we used the 2017-18 Multiple Indicator Cluster Survey (MICS) as the external source to obtain the target distributions for age groups (18-39 and 40+), education level (non-primary, secondary, and tertiary), a summary index of household asset ownership (based on whether the household had electricity, a durable roof, and access to an improved water source), household size (1-5, and 5+ members), and urban/rural residence. Because we worked in two provinces only, we calculated separate sets of weights for Kinshasa and North Kivu, and combined them to obtain final weights based on the proportion of the population in Kinshasa (63.5%) and North Kivu (36.5%) among the total population of these two provinces. To avoid high variability when using post-stratification weights, we limited the maximum weight to 2 in the three surveys (35).

#### 2.2.2 Imputation to obtain full SSH

As we did not know the age at the time of the interview for surviving siblings and the age at death for deceased siblings who died before 2019, we could not estimate mortality directly and had to resort to imputation or modelling. Building on previous work referring to child mortality and birth histories (36), we imputed ages and dates of death from an earlier DHS survey onto the summary sibling data collected in the MPS. We used the 2021 DHS in Burkina Faso, the 2015-16 DHS in Malawi, and the 2013-14 DHS in DRC.

We first imputed an age difference between each respondent and their surviving siblings. This age difference was sampled from distributions found in the DHS, tabulated for each five-year age group of respondent, and, in Burkina Faso and DRC, by sex of the sibling. From the age difference and the age of respondent, we recalculated the age at the time of the survey for all surviving siblings. For siblings who had died, we cross-tabulated the siblings’ age at death and time since death from the DHS, categorized by the five-year age group of respondents and, in Burkina Faso and DRC, by sex of the siblings. Based on these matrices, we proportionally sampled an age at death and a time since death for each deceased sibling. From the imputed values, we could then calculate the date of death and infer the date of birth for deceased siblings.

To estimate mortality, two approaches were then used, and we will refer to these as “partial” and “full” imputation. For the partial imputation, we calculated mortality from 2019 onwards, using the ages at death and dates of death of siblings who had died recently, as reported by the MPS respondents, and using the imputed data only to obtain an age for the surviving siblings. For the full imputation, we discarded all the information reported in the MPS on ages and dates for recent deaths, and used the imputed data instead

To ensure that our imputation approach produced acceptable results, we tested it on existing DHS where at least two surveys had included a module on sibling mortality (82 DHS from 41 countries, listed in Table S1). For each country, we retained the most recent DHS with this module and only retained the summary data as we had collected in the MPS. Based on sibling histories from a preceding DHS, we then applied the partial imputation (As described above, expect that we computed rates for the period 0-5 years before the survey to reduce sampling variability) and the full imputation. We calculated mortality rates from the imputed sibling histories, and compared the results with rates obtained directly from the reported ages and dates in the last survey. The results of this test are detailed in appendix A1. The full imputation provides results that are very close to the direct estimates: the mean absolute percentage deviation between the imputed and reference estimates is lower than 2.5% for each sex. The partial imputation is less precise and the relative deviations are quite large: the mean absolute percentage deviation is 23% for each sex. Still, in Burkina Faso, Malawi and DRC, the full imputation method gives acceptable results. We also have a relatively recent DHS to impute data from a full sibling history onto the age-specific distributions of total number of siblings and deceased siblings of the MPS.

### 2.4 Mortality estimation from MPS

After expanding the summary SSH collected in MPS to full SSH through imputation, we estimated adult mortality using a direct method, by dividing the deaths by the corresponding exposure times, by age and by period since data collection. We converted the mortality rates into survival probabilities, chained the survival probabilities together and converted these cumulative probabilities into _35_q_15_, the probability that an adolescent aged 15 years would die before reaching age 50 years. We used the "demogsurv" package of the statistical software R (https://github.com/mrc-ide/demogsurv). Confidence intervals around the probability _35_q_15_ were obtained through the jackknife method. The probability _35_q_15_ was retained as this is a common summary measure of adult mortality. It is adequate for synthesizing SSH data, as respondents are generally aged 15-49 (e.g., women in DHS) and the mean age of their siblings is typically close to their own age.

## 3. Results

### 3.1 Characteristics of the MPS samples

Fig 1 summarizes the information from Tables S6, S7, and S8 in the appendix that compare respondent characteristics to the general population in each country. The reference populations are based on the 2019-2020 census for Burkina Faso, the 2018 census for Malawi, and the 2017-2018 MICS survey for the DRC.

**Fig 1:**
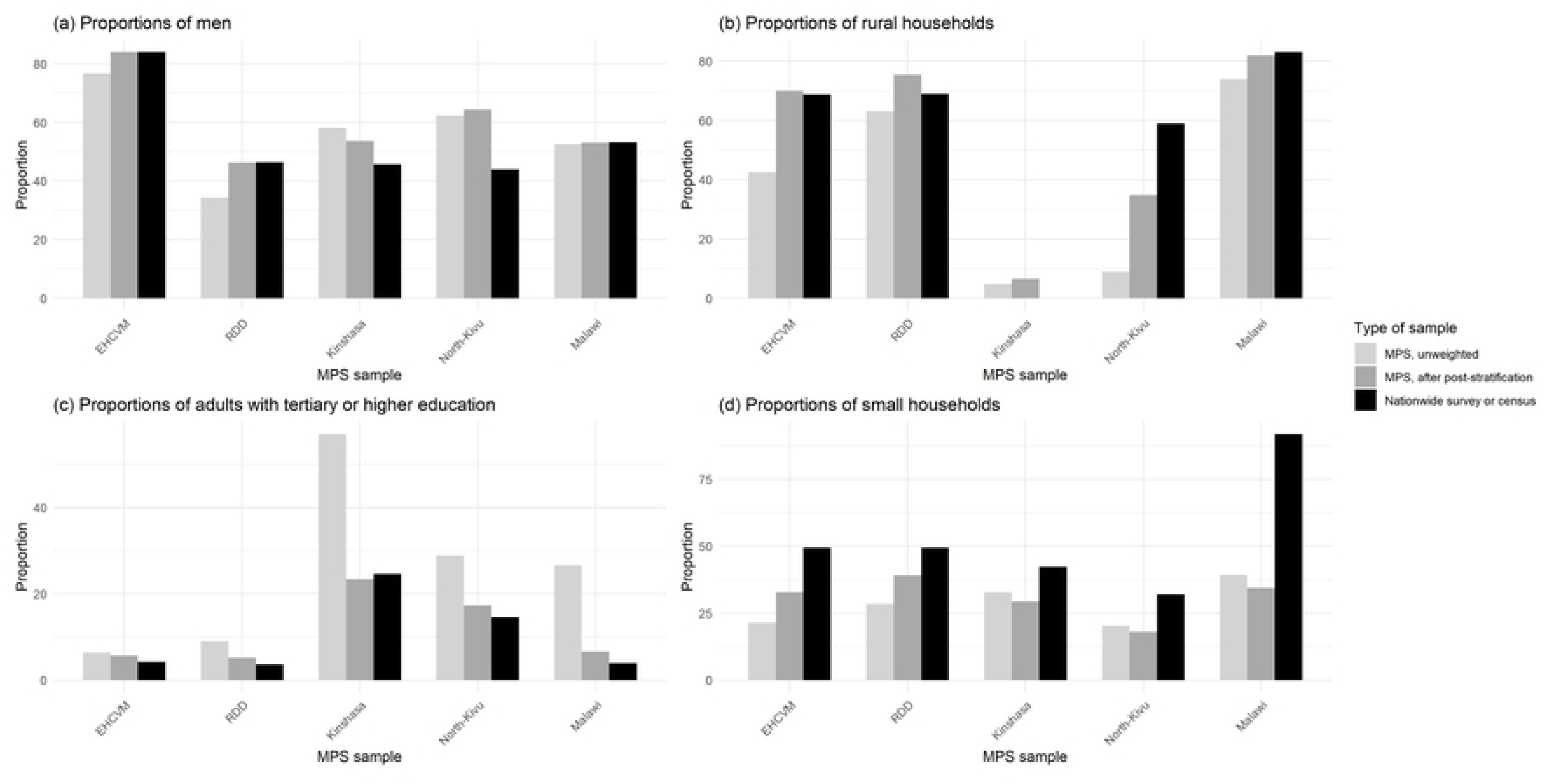
Composition of the MPS samples and comparison with nation-wide censuses or surveys in Burkina Faso, Malawi and DRC.

The characteristics of the MPS respondents differ systematically from those of the reference national populations. In the three MPS, there is an under-representation of rural respondents. For instance, in Burkina Faso, rural respondents constitute approximately 43% and 63% of the EHCVM and RDD samples, respectively, while they account for 69% of households in the census (Rural adults aged 15 to 64 also account for 69% of adults of the same age in the census). In Malawi, rural respondents represent 74% of the surveyed population in MPS, compared to 83% of the population enumerated in the 2018 census. The population with higher levels of education is also overrepresented in the MPS. Finally, all MPS are characterized by the underrepresentation of respondents living in small households. The difference is striking in Malawi, where 39% of MPS respondents live in households with 1-4 members, against 92% in the 2019 census.

After applying post-stratification weights, our three samples have characteristics that are comparable to our reference populations, with some exceptions. In all three surveys, members of larger households remain over-represented even after weighting. In the DRC, we also retain an over-representation of men, and of the urban population in North Kivu.

### 3.2 Assessment of the quality of data on summary sibling histories

To evaluate the quality of SSH, we first examined the mean number of reported siblings and compared each MPS with the last DHS survey that included a module on adult mortality (Fig 2). The number of siblings is expected to be slightly lower in the MPS than in the DHS conducted a few years earlier as a result of the fertility decline. In Burkina Faso, the two surveys were carried out only a year apart, but participants in the MPS tended to report a higher number of siblings, irrespective of the study arm. By contrast, in Malawi, the MPS shows the expected increase in mean number of siblings as the age of respondents rise, but with fewer siblings reported than in the 2015 DHS (up to one less sibling in the MPS in respondents aged 15-19). In DRC, there is virtually no difference between the two data sources in Kinshasa, but in North Kivu, respondents in the MPS, particularly the older age groups, reported slightly more siblings. However, these differences remain relatively small, averaging less than 0.5 siblings.

**Fig 2.**
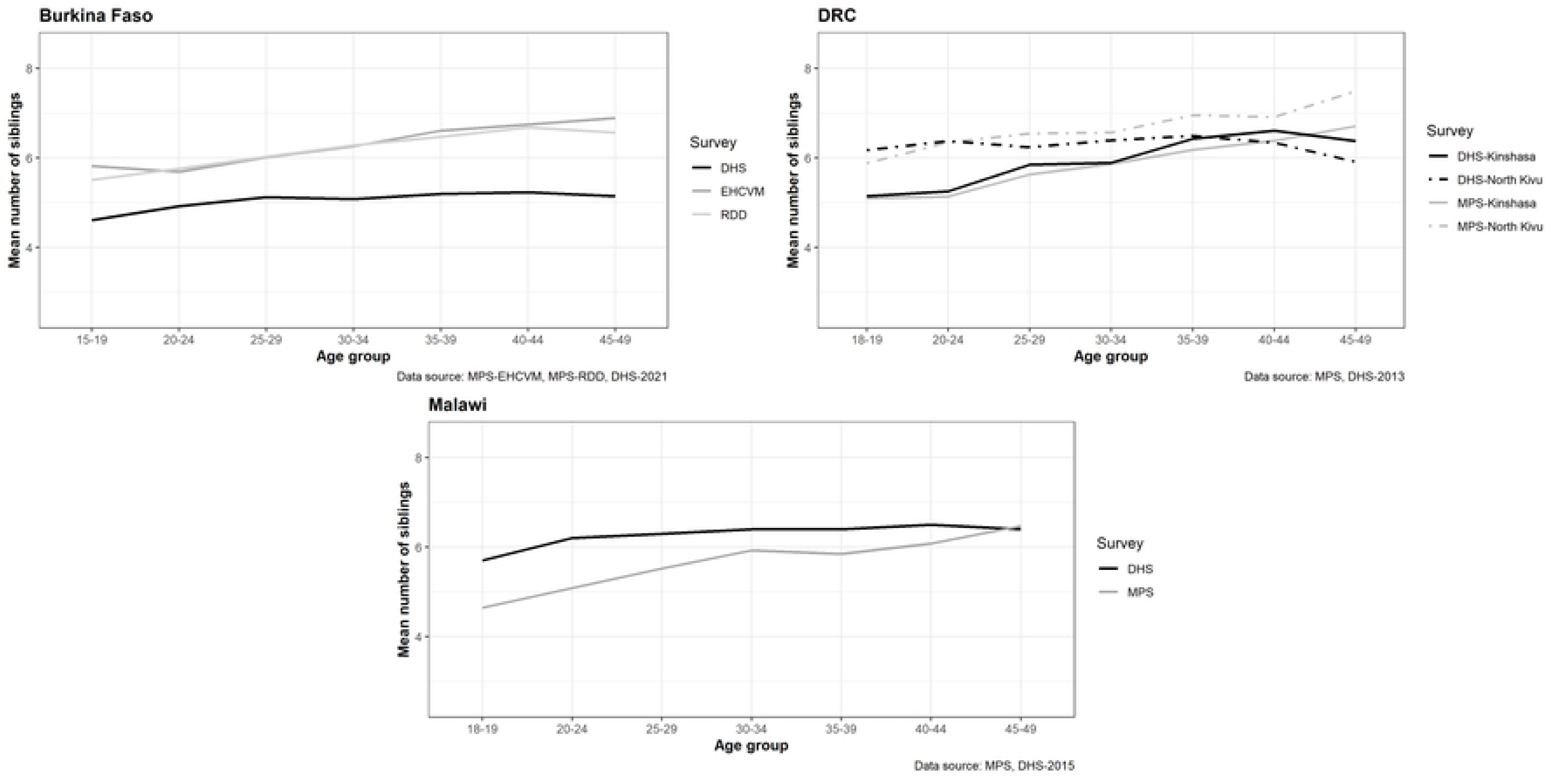
Mean number of maternal siblings by age group of respondent in the MPS and DHS conducted in Burkina Faso, Malawi and DRC. Note: In Malawi and the DRC, the MPS samples were restricted to ages 18 and above, and we also excluded respondents aged 15-17 in the DHS samples.

In Fig 3, we present the proportions of surviving siblings categorized by the age group of respondents in each MPS, again comparing with the latest DHS with full sibling histories (see Fig S4 for sex-specific results). As expected, there is a consistent decline in the proportion of surviving siblings as the age of respondent increases, regardless of the data source. However, there are significant differences between the data series, and inconsistent patterns in the three countries. In Burkina Faso, proportions of surviving siblings from both MPS study arms are lower than in the 2021 DHS, while the reverse is true in DRC in both provinces. In Malawi, there is a good agreement between the MPS and the 2015 DHS.

**Fig 3.**
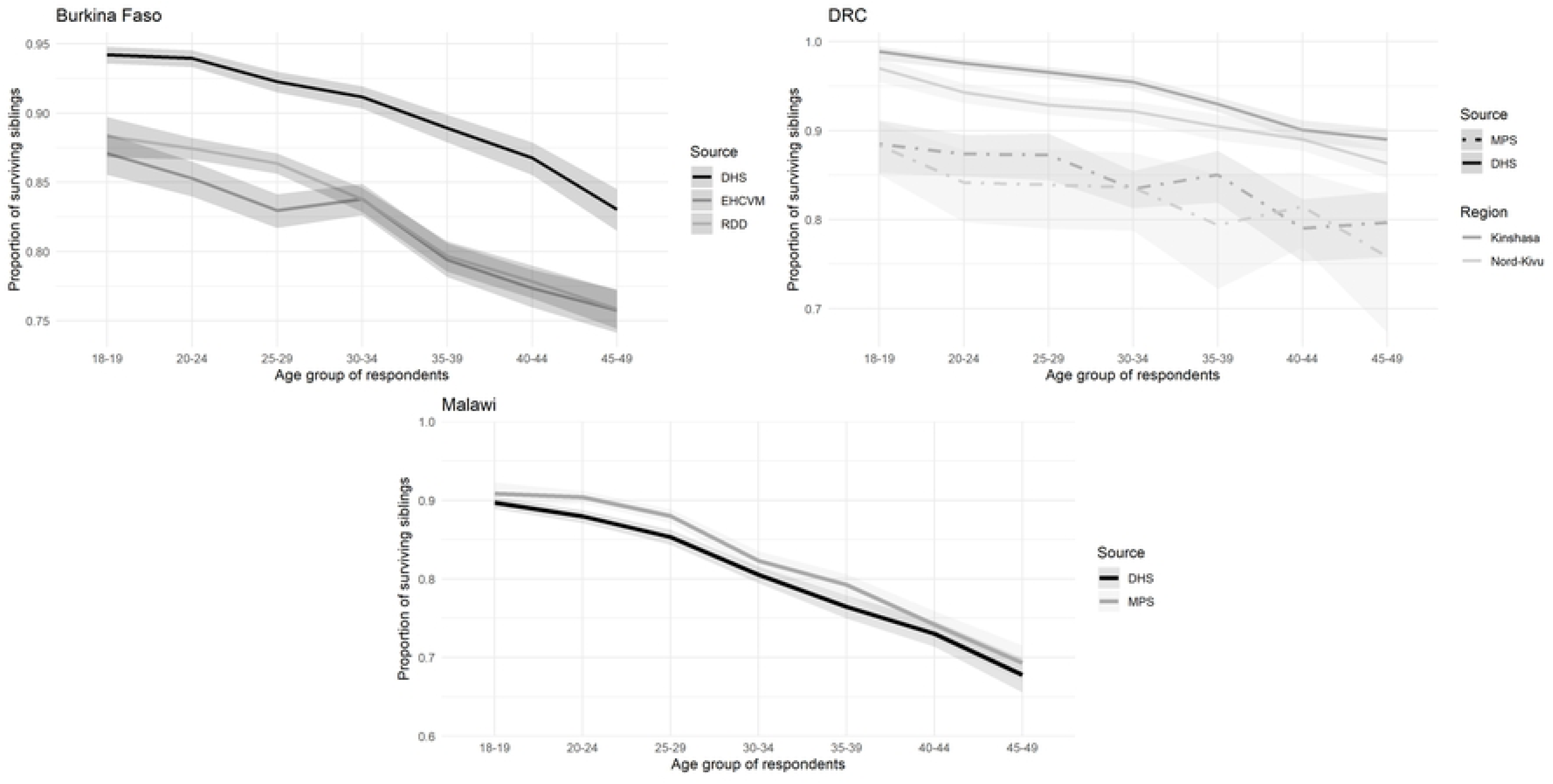
Proportions of surviving siblings at the time of the survey, by age group of respondent in MPS and DHS, in the three countries.

Fig 4 illustrates the proportion of deaths that occurred in the three years preceding the survey, classified by age group of the respondent. We compare these proportions to those calculated in the most recent DHS (see Fig S5 for sex-specific results). Regardless of the country, the proportions of recent deaths among siblings who have passed away are lower in the MPS than in the DHS, except for sisters in the Kinshasa province. While these disparities are relatively small in absolute terms, they have important implications for estimating recent mortality.

**Fig 4.**
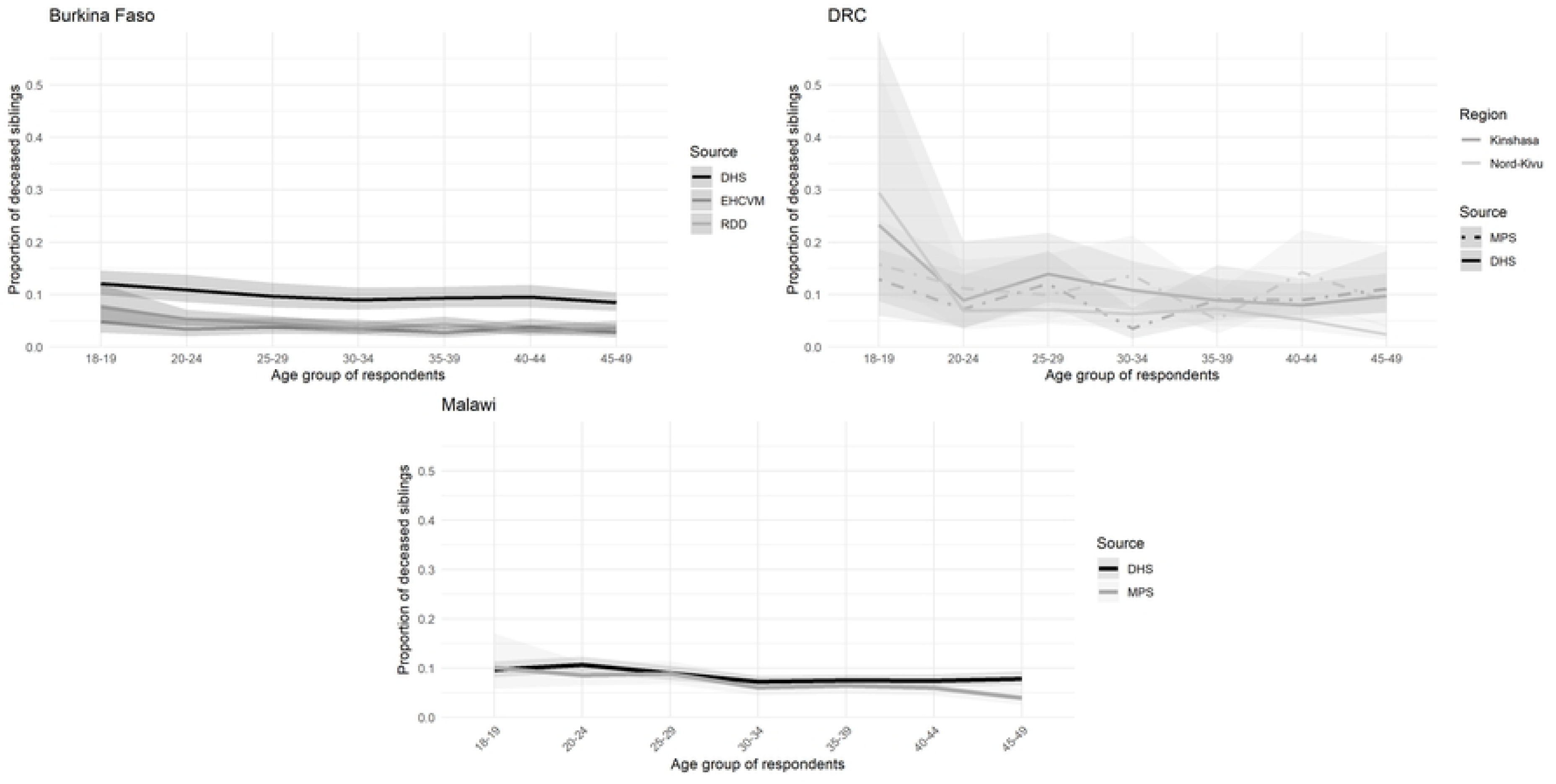
Proportions of recent deaths among all deceased siblings by age group and country (last 3 years in DHS and since January 2019 in MPS)

### 3.3 Adult mortality rates in MPS based on partial imputation

Table 2 presents the synthetic cohort probability that a 15-year-old adolescent dies before reaching age 50 (_35_q_15_). To put the MPS estimates in perspective, we compare them to those extracted from the most recent DHS. The DHS estimates are based on full sibling histories and are also calculated for the 0-3 year period preceding data collection. We contrast the survey data with estimates from the WPP, using the 2022 Revision. These draw on a multitude of sources, including sibling survival data, parental survival information, recent household deaths, and model age patterns of mortality [37] (The WPP estimates are available at the national level only, and to obtain reference adult mortality rates for DRC for the two provinces, we scaled the national adult mortality rates in the WPP according to the ratio of province-specific to national under-five mortality in the latest DHS. See appendix A.2 for more details). It is worth noting that the WPP estimates are usually higher than estimates from SSH collected in surveys, and this could be due to underestimation of mortality in SSH, or, in some countries, an upward bias in the WPP (38).

**Table 2:**
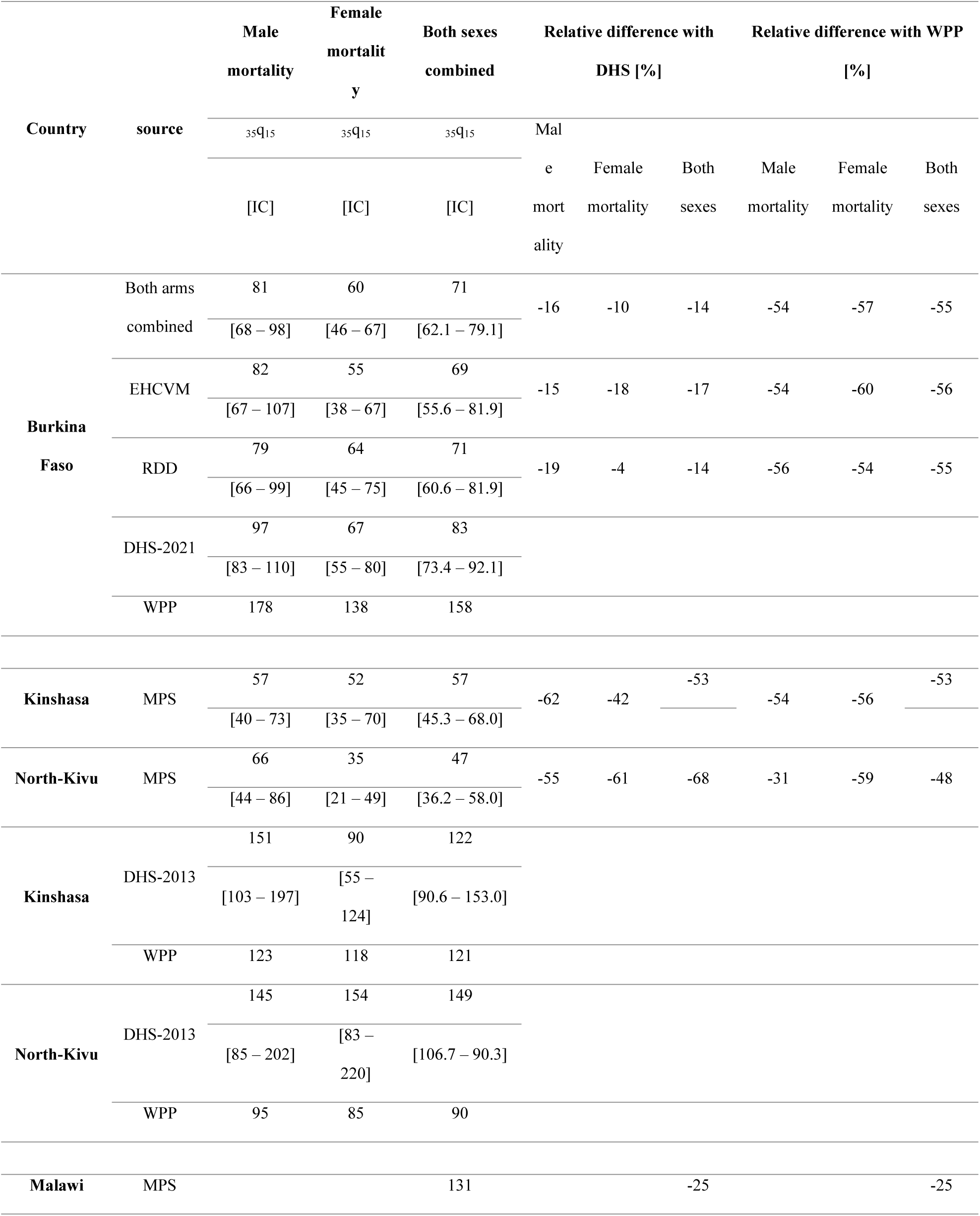
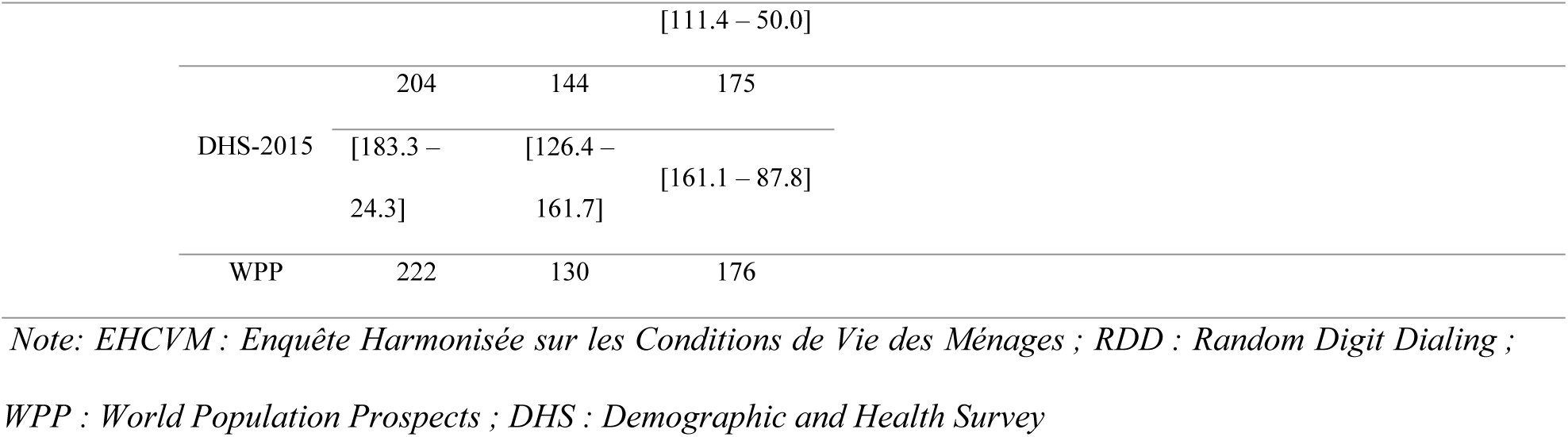
Estimates of the probability _35_q_15_ over the period 0-3 years before data collection, by source and country [‱], using partial imputation for the MPS [deaths before age 50 for 1000 adolescents aged 15].

Overall, the MPS estimates of adult mortality are lower than those in the latest DHS. In Burkina Faso, the MPS estimate of _35_q_15_ (both sexes) is 14% lower. In Malawi, _35_q_15_ is 25% lower than the estimate extracted from the 2015 DHS. It is, however, possible that adult mortality declined rapidly in-between the two surveys, for example thanks to the scale-up of antiretroviral therapy. In DRC, the differences with the pervious DHS are much larger (-53% in Kinshasa and -68% in North Kivu), but in this country, the comparison is more complicated as estimates refer to subnational areas.

MPS estimates are also considerably lower than those reported in the WPP. In Burkina Faso, for example, the WPP estimate of the probability that a 15-year-old would die before their 50th birthday in 2019 was 178 and 138 per 1,000 for men and women, respectively. In contrast, the MPS yielded estimates of 81 and 60 per 1,000 for men and women. This translates into a downward bias in the _35_q_15_ probability of 54% for men and 57% for women. The deviations are similar in the Kinshasa province of DRC (-54% for male mortality and -56% for female mortality), and slightly smaller in North Kivu (-31% and -59%, respectively). In Malawi, there is a better agreement across sources but the _35_q_15_ estimate for both sexes combined is still 25% lower than the corresponding probability in the WPP.

Annual estimates in the probability _35_q_15_ are also presented in appendix (Fig S6), showing fluctuations in mortality, but no evidence of a spike in mortality associated with the COVID-19 pandemic.

In Burkina Faso, the sex of the respondent or the sex of siblings do not appear to be systematically associated with deviations from the most recent DHS or WPP. For example, in the RDD arm, male respondents provide lower estimates than female respondents, but the reverse is true in the EHCVM arm [Table S9]. No systematic pattern emerges in the DRC either. In Malawi, mortality rates extracted from male respondents are 37% higher than based on female respondents, but the difference is not significant. An earlier study comparing reports on sibling survival according to the sex of respondents in 10 DHS did not detect significant or systematic differences in mortality estimates obtained from men and women either [37]

### 3.4 Adult mortality rates in MPS based on full imputation

Disparities in adult mortality levels observed between the MPS (using the shortened questionnaire) and the DHS (based on full sibling histories) could be caused by misreporting ages and dates of death. To address this, we imputed both ages and dates of death of siblings and compare those to the DHS and WPP Table 3 presents the synthetic cohort probabilities that a 15-year-old individual will die before reaching the age of 50 (_35_q_15_), based on the full imputation, comparing the MPS values with those from the most recent DHS surveys and the WPP. Mortality trends are displayed in Fig 5.

**Fig 5:**
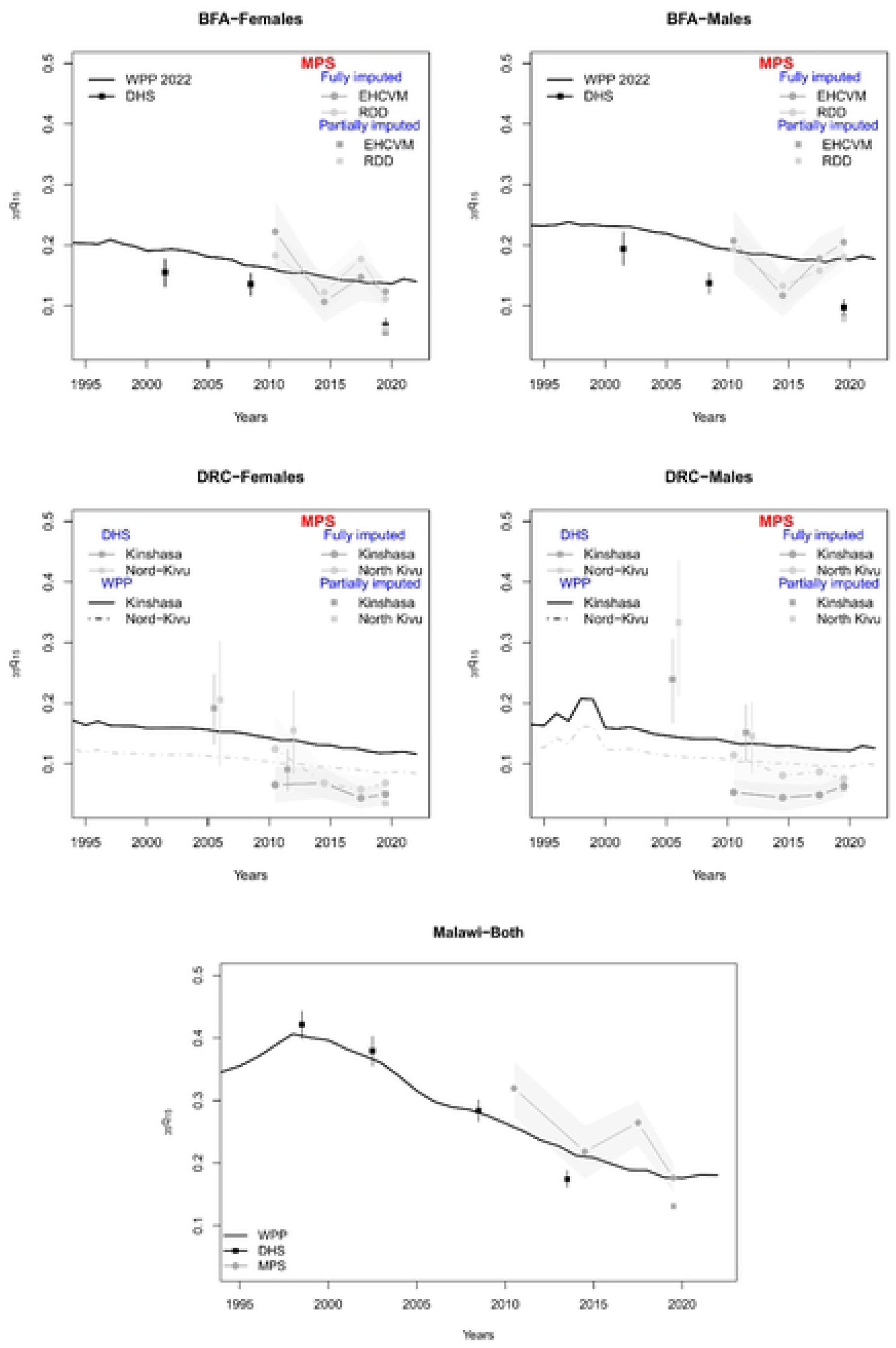
Trends in adult mortality [_35_q_15_] according to SSH in the MPS survey or DHS surveys and in the World Population Prospects.

**Table 3:**
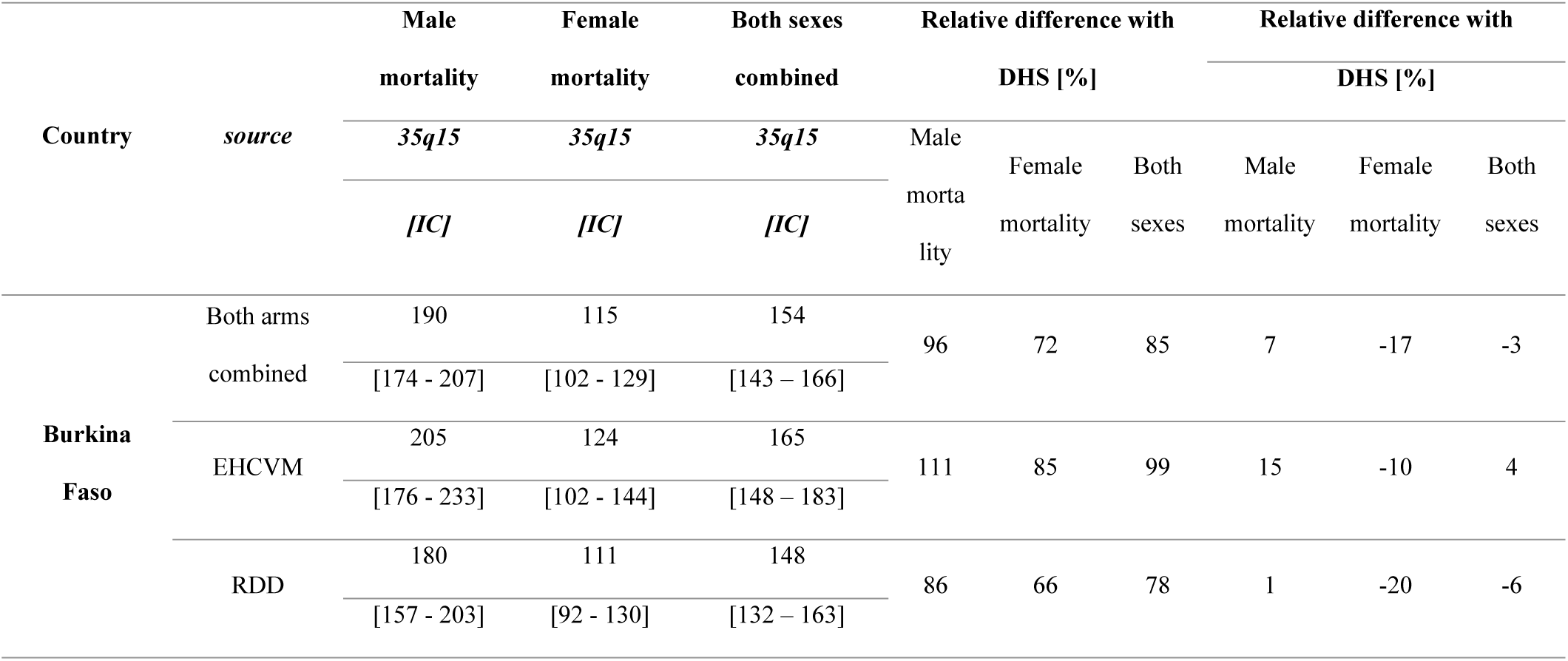
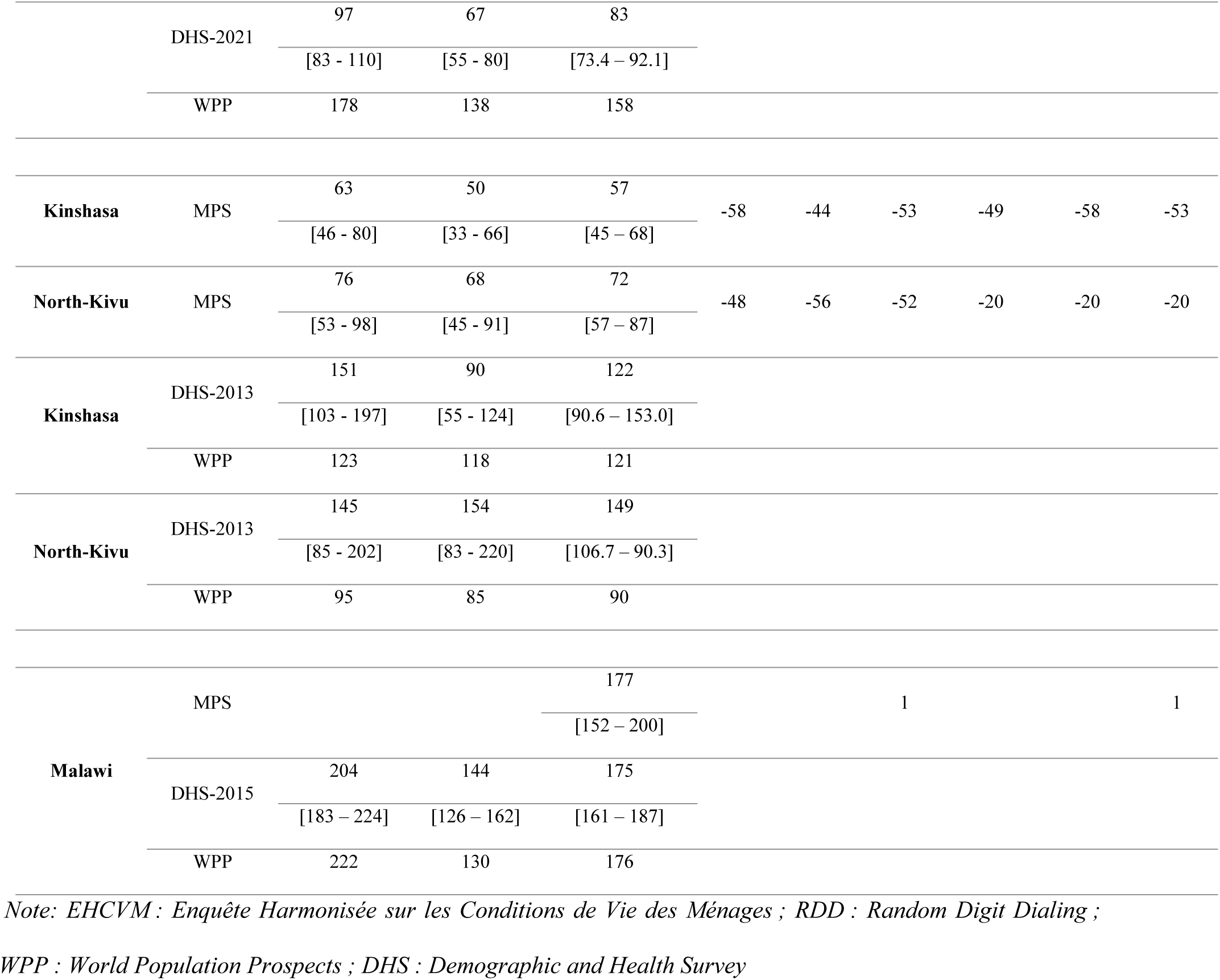
Estimates of the probability _35_q_15_ over the period 0-3 years before data collection, by source and country [‱], using full imputation for the MPS [deaths before age 50 for 1000 adolescents aged 15].

The MPS estimates obtained after full imputation are higher than those based on the partially imputed data pertaining three years. In Burkina Faso, the _35_q_15_ estimate from the MPS is now 85% higher than extracted from the 2021 DHS, while in Malawi, our more recent estimate is virtually the same as the probability estimated from the DHS 2015 (+1%). By contrast, in the DRC, they remain lower than the 2013 DHS, with relative differences exceeding 50%, regardless of the province.

MPS estimates based on the full imputation in Burkina Faso and Malawi closely align with those from the WPP. For instance, in Burkina Faso in 2019, the WPP estimated that, out of 1,000 individuals aged 15, 158 would die before reaching age 50, while the corresponding MPS estimate is 154 per 1,000 (-3% only). In Malawi, the relative difference is only 1% for both sexes combined. In contrast, in the DRC, regardless of the province, MPS estimates are still well below the reference values from the WPP (-20% and -53% for North Kivu and Kinshasa, respectively).

A breakdown by study arm in Burkina Faso shows that the EHCVM provided slightly higher estimates [165‱ [95% CI: 148 – 183], compared to 148‱ [132 – 163] in the RDD arm, see Table S10]. A breakdown by the sex of respondent shows that in Burkina Faso and Malawi, estimates based on the full imputation are higher when obtained from female respondents than from male respondents, regardless of the gender of the siblings. In the DRC, no systematic pattern emerges according to the sex of respondents.

## 4. Discussion

In this study, we compared adult mortality from MPS in Burkina Faso, Malawi and the DRC with estimates from the DHS and the WPP.

In comparison to the DHS Sibling Survival Histories, the MPS included a shortened version of the set of questions, and we developed two imputation approaches to reconstruct full sibling histories. To assess the validity of these imputation approaches, we first tested them on existing data from 41 DHS. Our results suggest that only imputing the ages of surviving siblings from an earlier survey yields unbiased estimates. In our test with existing DHS, all the imputed estimates fell within the 95% confidence intervals around the direct estimates obtained from the reported ages. In contrast, imputing all ages and dates yielded less precise estimates, especially when surveys were spaced far apart, or, in countries with rapididly changing mortality (e.g., due to HIV epidemic and the expansion of treatment programmes). This is because this approach assumes that the distributions of age differences and the timing of deaths that prevailed in the past still apply in the more recent period. This imputation approach is also likely to produce imprecise results when there are differences in the sample composition of successive surveys.

All three MPS underestimated adult mortality levels when relying on all dates and ages gathered over the phone, but the extent of this underestimation is difficult to establish as there is considerable uncertainty surrounding expected mortality rates among adults in these countries. However, it is likely that the MPS underestimated adult mortality, because the rates based on all reported dates and ages were still lower than those extracted from full sibling histories collected in face-to-face interviews. This could be a consequence of the use of a shorter series of questions, offering fewer opportunities to correct statements, and giving rise to more confusion among respondents. This could have led to omissions of deaths or transfers of some recent deaths out of the reference period (2019-2022). We observed that the MPS registered a much lower proportion of recent deaths than in the DHS. It is worth noting that supplementary questions were asked for recent deaths, such as those related to the circumstances of death. This might have prompted interviewers to move certain deaths out of the reference period to reduce their workload [38]. Respondents themselves could face difficulties in dating recent sibling deaths when sibling histories take the form of the small set of questions used in the MPS. Indeed, when we compared the summary sibling data in the MPS and the DHS, we observed only small discrepancies between the two sources, when assessing sibship sizes and the proportions of surviving siblings. Furthermore, the adult mortality estimates obtained after imputing all dates of birth and death were closer to the estimates from previous DHS or the WPP estimates than those calculated using the reported ages and dates for recent deaths.

It is possible that these differences are in part attributable to the mode of collection, with errors more frequent by telephone than face-to-face. The low mortality rates recorded in MPS could also be the result of selection biases related to the characteristics of mobile phone users. Indeed, our analyses revealed several significant differences between the composition of the phone survey samples and those of previous face-to-face surveys. Respondents living in rural areas and those with lower educational levels were underrepresented in our MPS samples, particularly in DRC, where the difference was significant. A more detailed comparison, not presented here, between participants in the MPS in Burkina Faso and all household heads who owned a mobile phone in the original face-to-face survey conducted in 2018-2019 (EHCVM) also showed that phone survey participants were generally younger, more educated, more frequently resided in urban areas, and lived in larger households, and lived in larger households with more amenities [39]. These findings align with recent work in other low- and middle-income countries [7,41]. While we accounted for these biases through post stratification weighting, it is possible that our weighting adjustments do not entirely remove all selection biases. A recent study based on mobile phone owners in DHS suggested that post-stratification weighting of sibling histories based on respondents’ background characteristics did not always produce a correction in the expected direction and needs to be used with caution [42]. Detailed sociodemographic information about customers using various cell phone networks could help investigate theses errors further. For future MPS, addressing potential under-representation can be achieved through the implementation of an adjusted quota sampling strategy, contingent on the availability of sociodemographic and economic data on mobile phone network customers.

Despite these limitations, our analyses of mortality levels and trends in Burkina Faso, Malawi, and DRC reveal the potential to conduct large-scale MPS for collecting mortality data. In Burkina Faso and Malawi, adult mortality trends derived from the MPS aligned relatively well with estimates from the United Nations, when we used the full imputation approach. More research is needed on the exact set of questions and their wording to collect shortened summary data, in cases where the collection of full sibling histories is not possible. For example, asking about the total number of siblings who have reached age 15, those who have died and how many of these adult deaths occurred recently makes it possible to apply indirect estimation [43,44]. Future studies should evaluate the quality of the data collected on summary sibling histories, by randomly assigning respondents to different versions of the survey instrument. Future MPS should also pilot the collection of full sibling histories to better evaluate the potential of this module to be associated with other interview modalities than face-to-face surveys. Additionally, an effective communication strategy informing the population about the data collection period and the objectives of the MPS could play a role in reducing interview time and enhancing respondent cooperation, thereby significantly enhancing the quality of the data collected.

## Declarations

### Ethics approval and consent to participate

The study protocol for Burkina Faso was reviewed by Ethics Committee of the Ministry of Health of Burkina Faso, Ethics Committee of the London School of Hygiene and Tropical Medicine (reference: 26393/RR/24486) for DRC, and the University of Malawi Research Ethics Committee (reference: P.07/21/76) for Malawi. All participants provided oral consent for the survey, including consent for storing anonymized data in a public repository and consent to audio record the interview.

### Availability of data and materials

RaMMPS data can be requested via email to the corresponding author or.

### Authors’ contributions

KD, SL and BM analyzed the MPS and DHS data. All authors read and approved the final version of the manuscript.

## Appendix

## A.1. Evaluation of the imputation approach on existing DHS in Sub-Saharan Africa

This section presents results related to the imputation method applied to existing DHS.

Fig S1 depicts the male probability _35_q_15_ estimated directly from the full SSH in the latest DHS, compared to the probability obtained when we impute the ages of surviving siblings from an earlier survey in the same country. The differences between the two sets of estimates are negligible [Table S1]. Absolute differences vary from -5‱ in Guinea to 14‱ in Togo (for females). Relative differences vary from -3% in several surveys to 10% in Morocco (for men), and the mean absolute percentage deviation between the imputed and reference estimates is lower than 2.5% for each sex. In all DHS, the imputed estimate falls within the 95% confidence intervals around the direct estimate obtained from the reported ages.

In contrast, there are relatively large deviations between estimates calculated directly and those based on full imputation (Fig S2). The median ratio of imputed to reference estimates is 0.89 for men and 0.96 for women, indicating that this approach tends to produce under-estimates, except for a few countries. Absolute differences range from -85‱ in Haiti to +138‱ in Zimbabwe (for men). Even if we account for the fact that both the reference and imputed estimates are affected by sampling errors, the relative deviations are quite large: the mean absolute percentage deviation is 23% for each sex. For male mortality, the estimates based on imputation falls outside of the confidence intervals around the reference estimates in 27 surveys, representing about two thirds of our sample. This is the case in 30 surveys for female mortality. There are several potential explanations for these discrepancies. Firstly, some surveys may have occurred at intervals that are too far apart, to the point where the structure of sibships or the level and age pattern of mortality of the previous survey no longer accurately reflects the experience captured in the more recent survey. This could be the case in South Africa, for example, where there is a gap of 20 years between successive surveys. In other countries, the disruptions introduced by the HIV/AIDS epidemic could play a role in these discrepancies; the full imputation works particularly badly in Namibia or Zimbabwe, where adult mortality has declined thanks to the scale-up of ART. It is also possible that differences between surveys in terms of data quality or sample composition lead to such deviations. Comparisons of the mean number of siblings reported per cohort of respondents are not always consistent, and in some cases, the numbers reported in the most recent survey are implausibly low, when compared to the preceding survey (Fig S3).

Despite these limitations, the imputation method produces acceptable results in several countries. In the three countries in which we carried out the MPS, the mean absolute percentage deviation averages 17.7% for men and 12.7% for women in the DHS. In these three cases, we also have a relatively recent DHS to impute data from a full sibling history onto the age-specific distributions of total number of siblings and deceased siblings of the MPS to generate mortality estimates.

**Fig S1:**
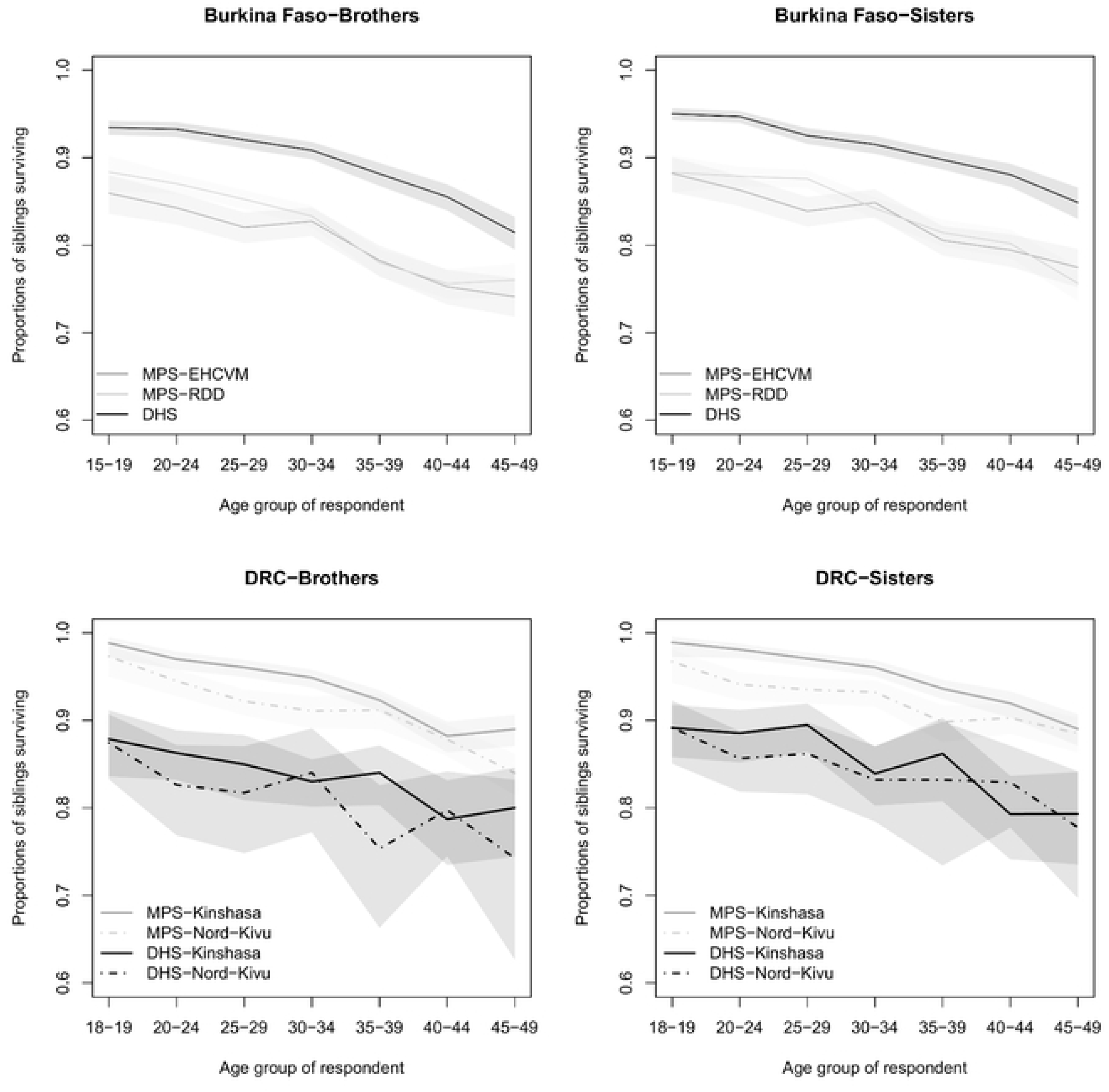
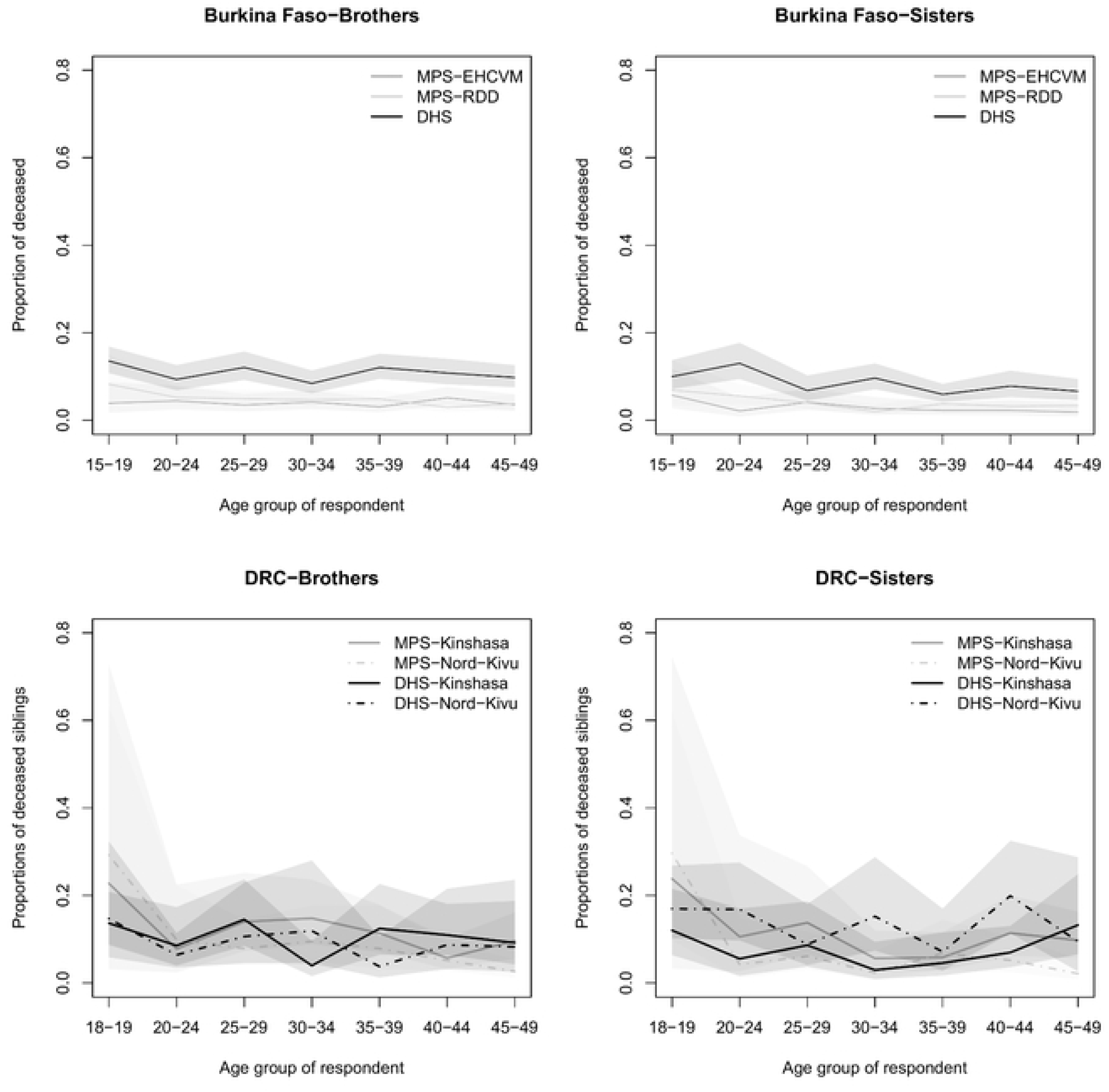
Risk of dying in adulthood (_35_q_15_) in females, as obtained from full SSH in the latest DHS with sibling data (0-5 years prior to the survey), compared to levels based when imputing the ages of surviving siblings from a preceding DHS.

**Fig S2:**
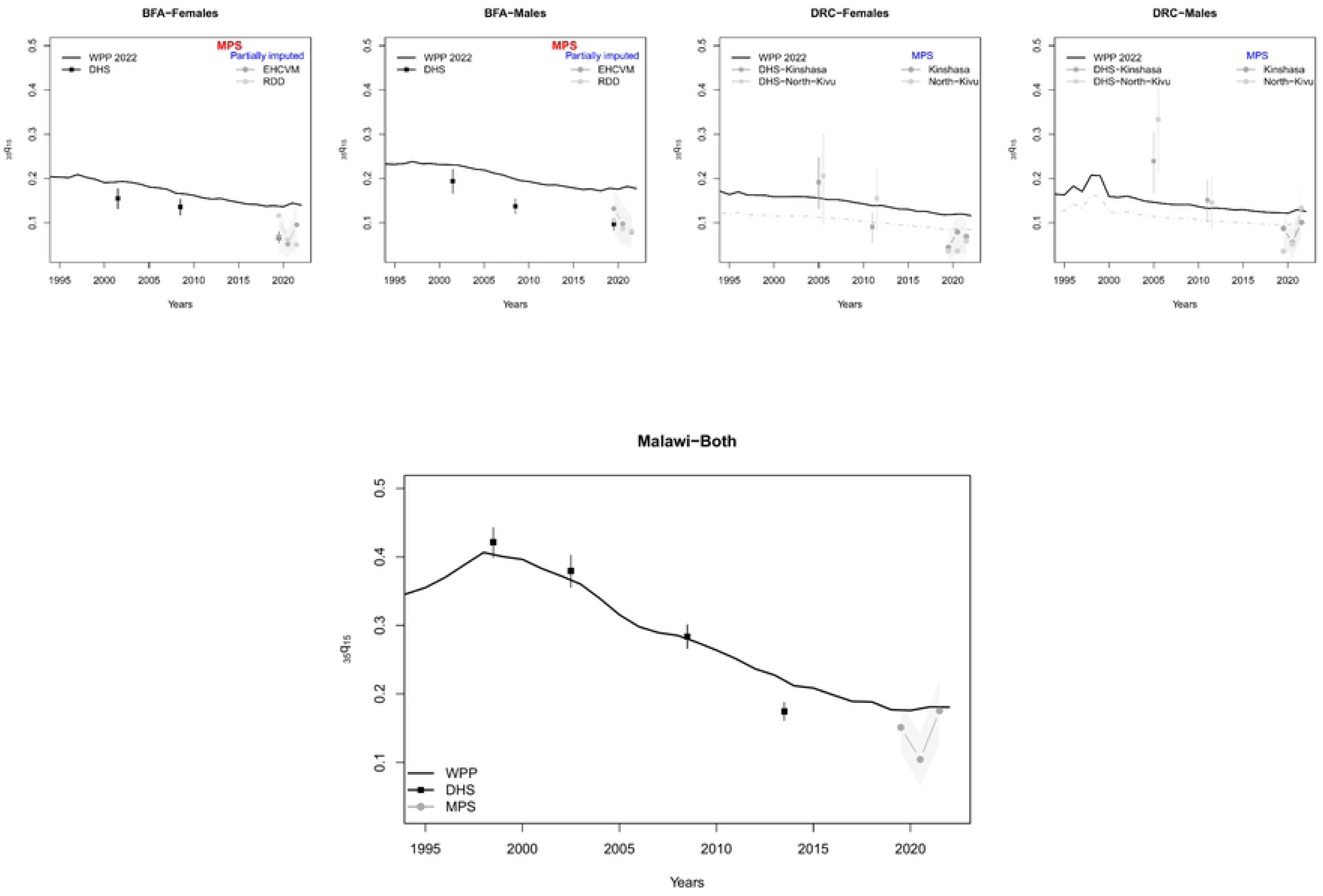
Risk of dying in adulthood (_35_q_15_) in females, as obtained from full SSH in the latest DHS with sibling data (0-5 years prior to the survey), compared to levels based when imputing all ages and dates from a preceding DHS.

**Table S1:**
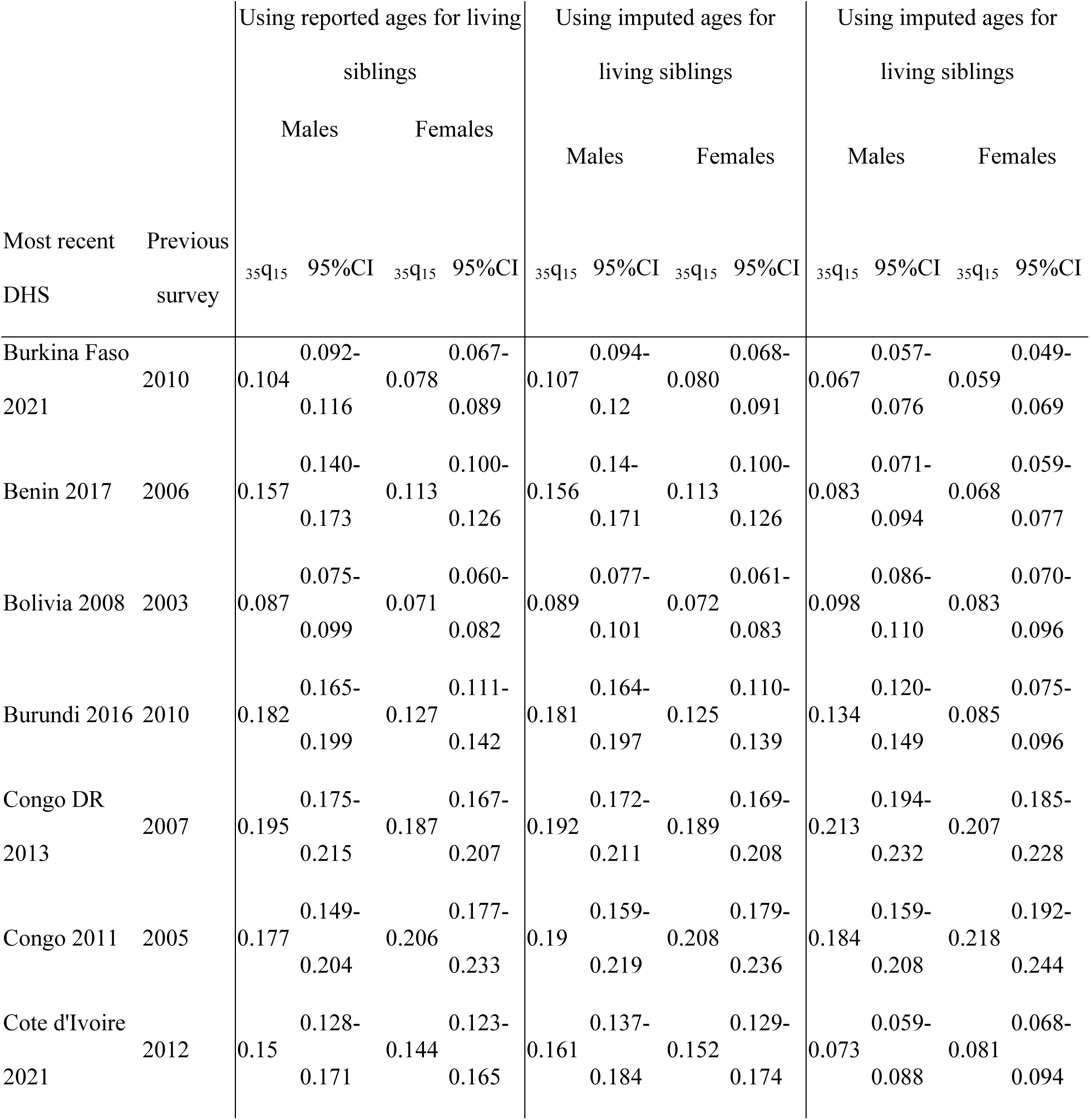
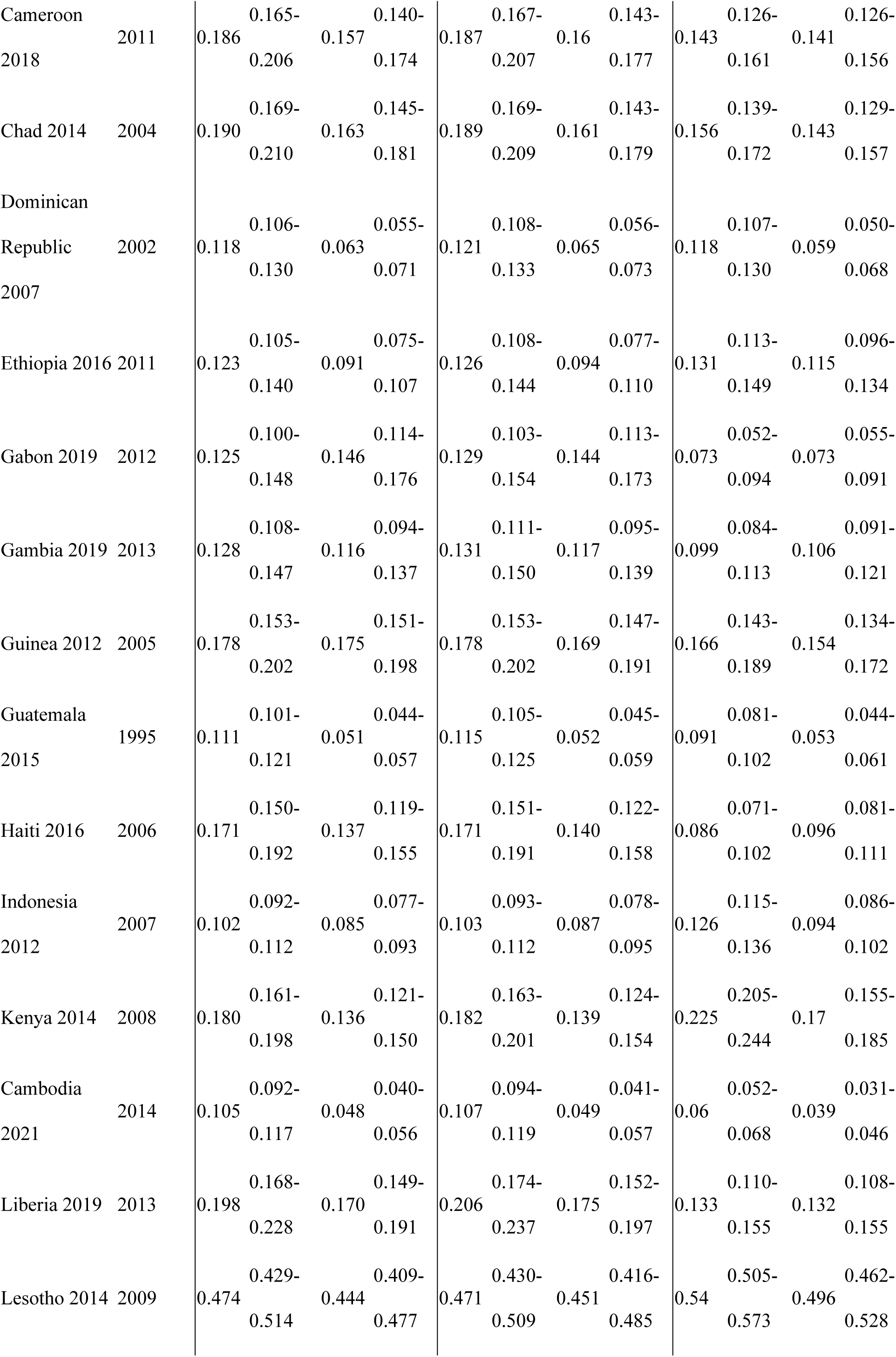
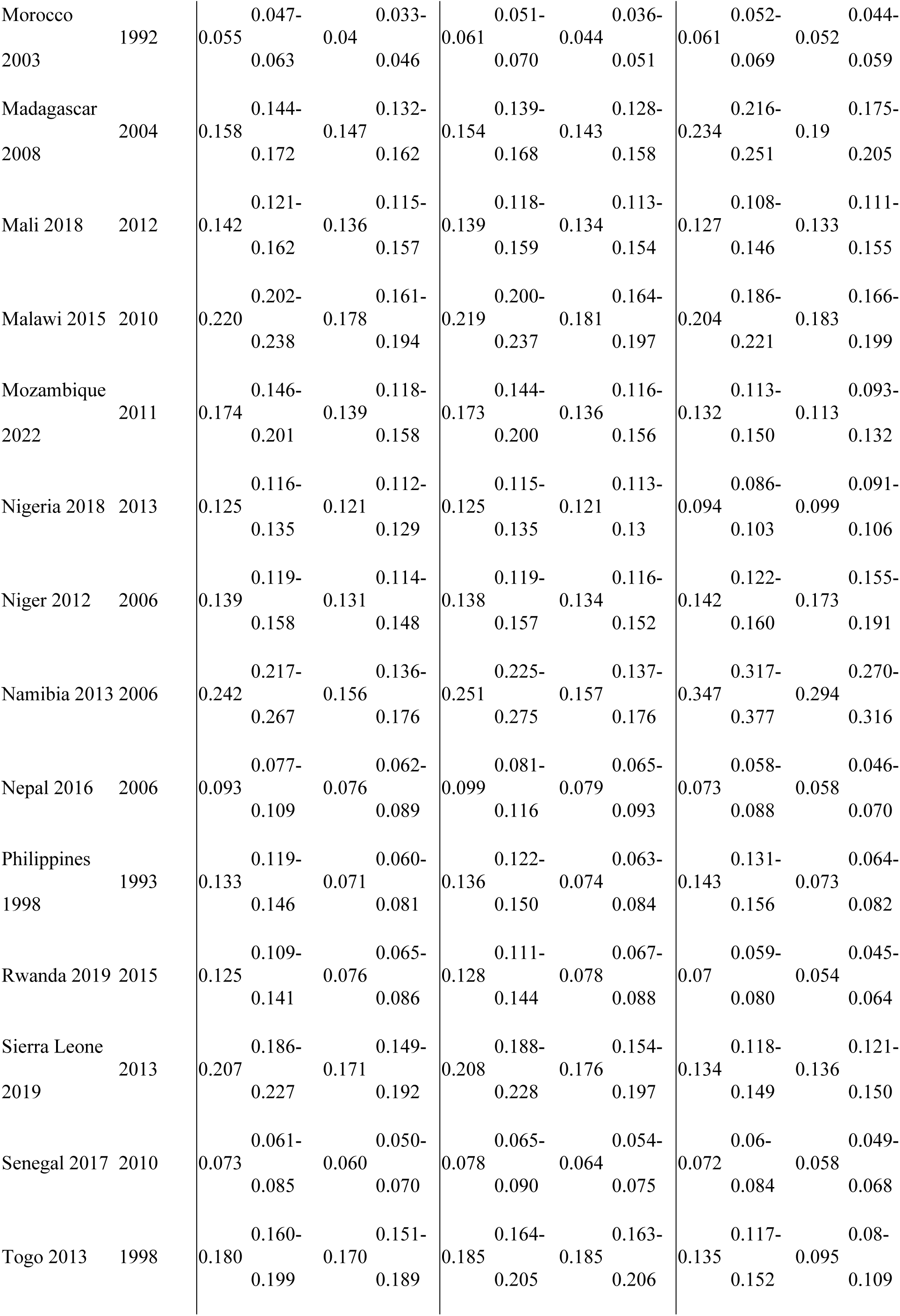
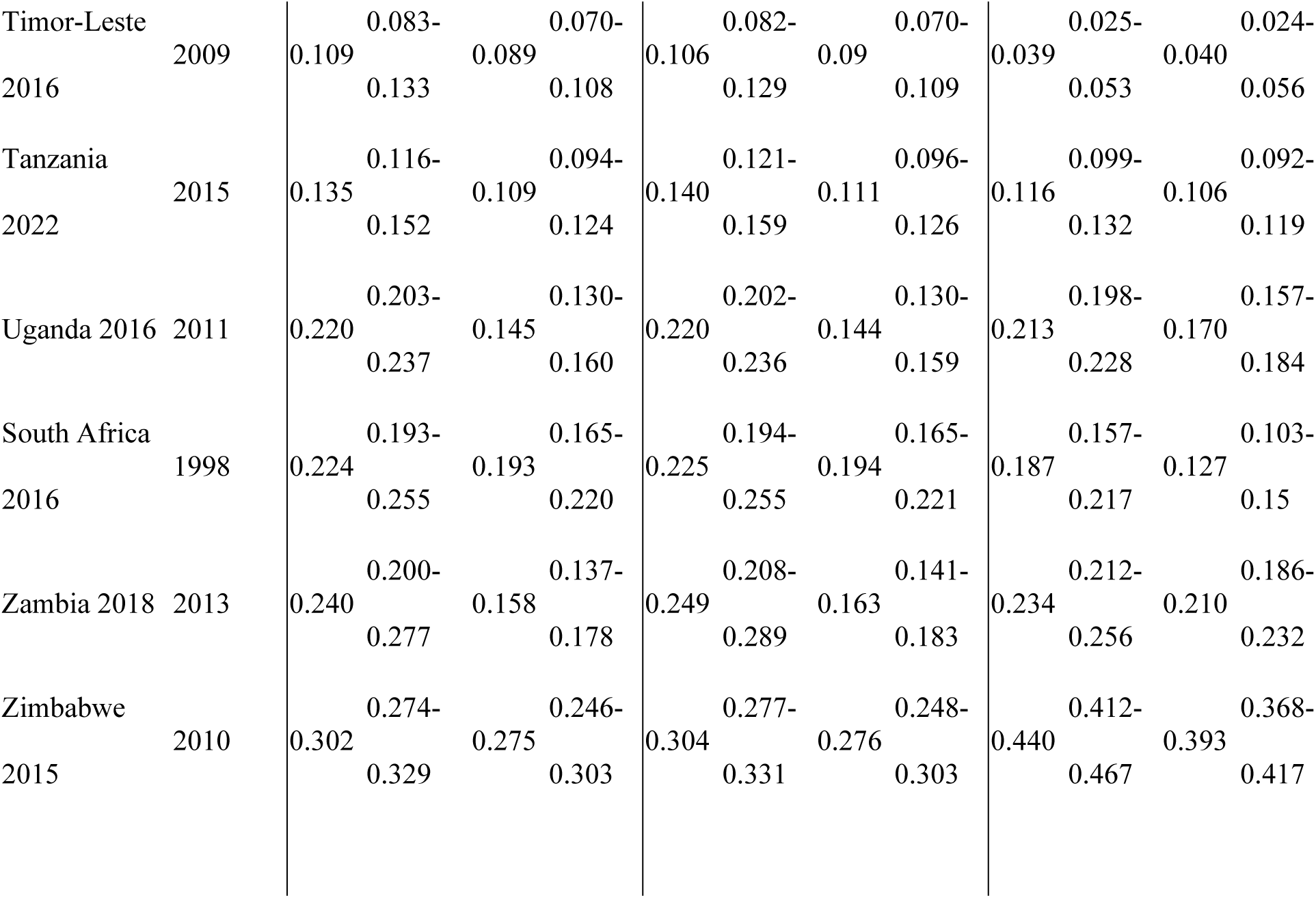
Estimates of the probability _35_q_15_ estimated from the most recent DHS with sibling histories, without imputation and with imputation based on a previous DHS.

**Fig S3:**
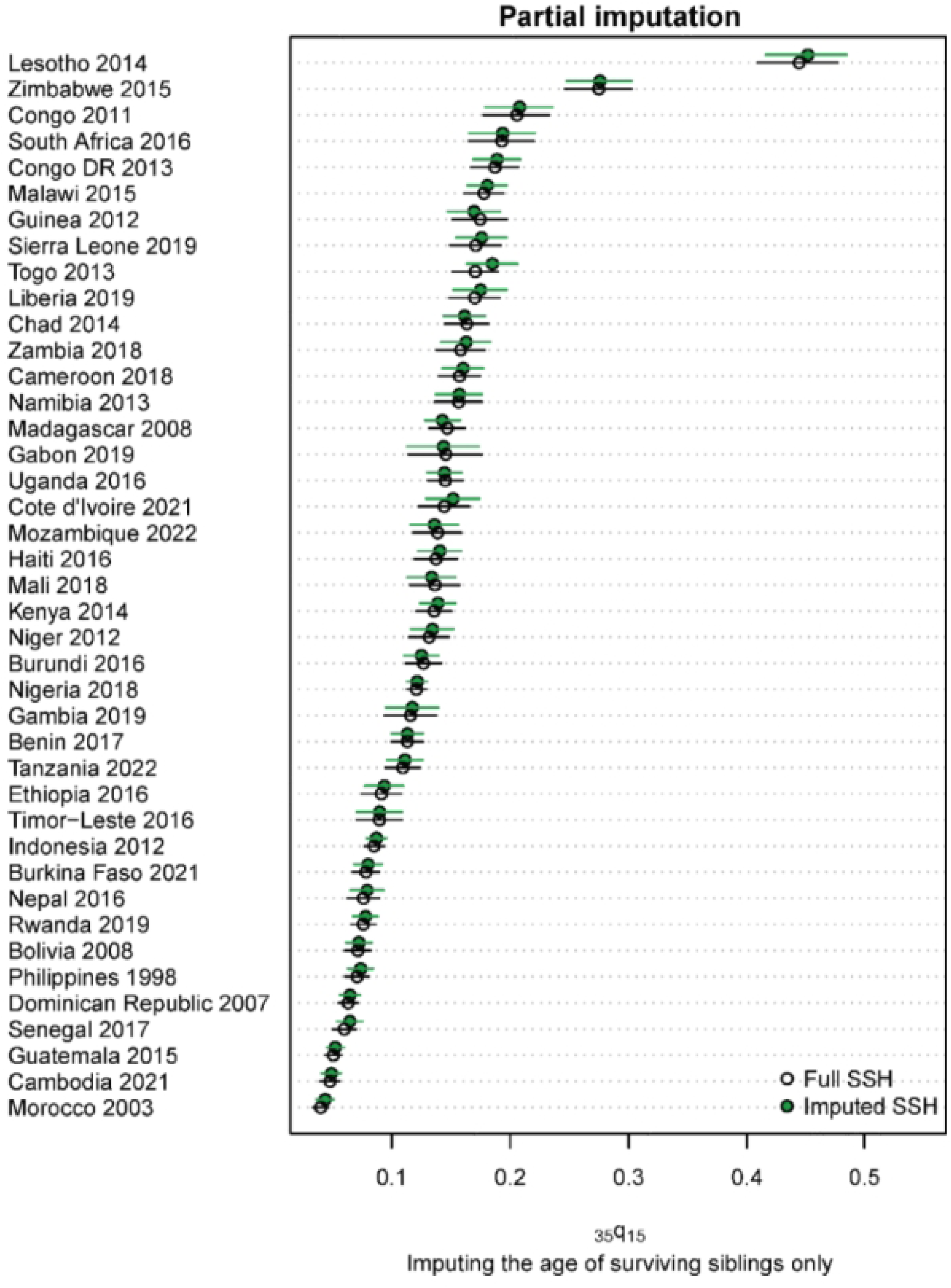
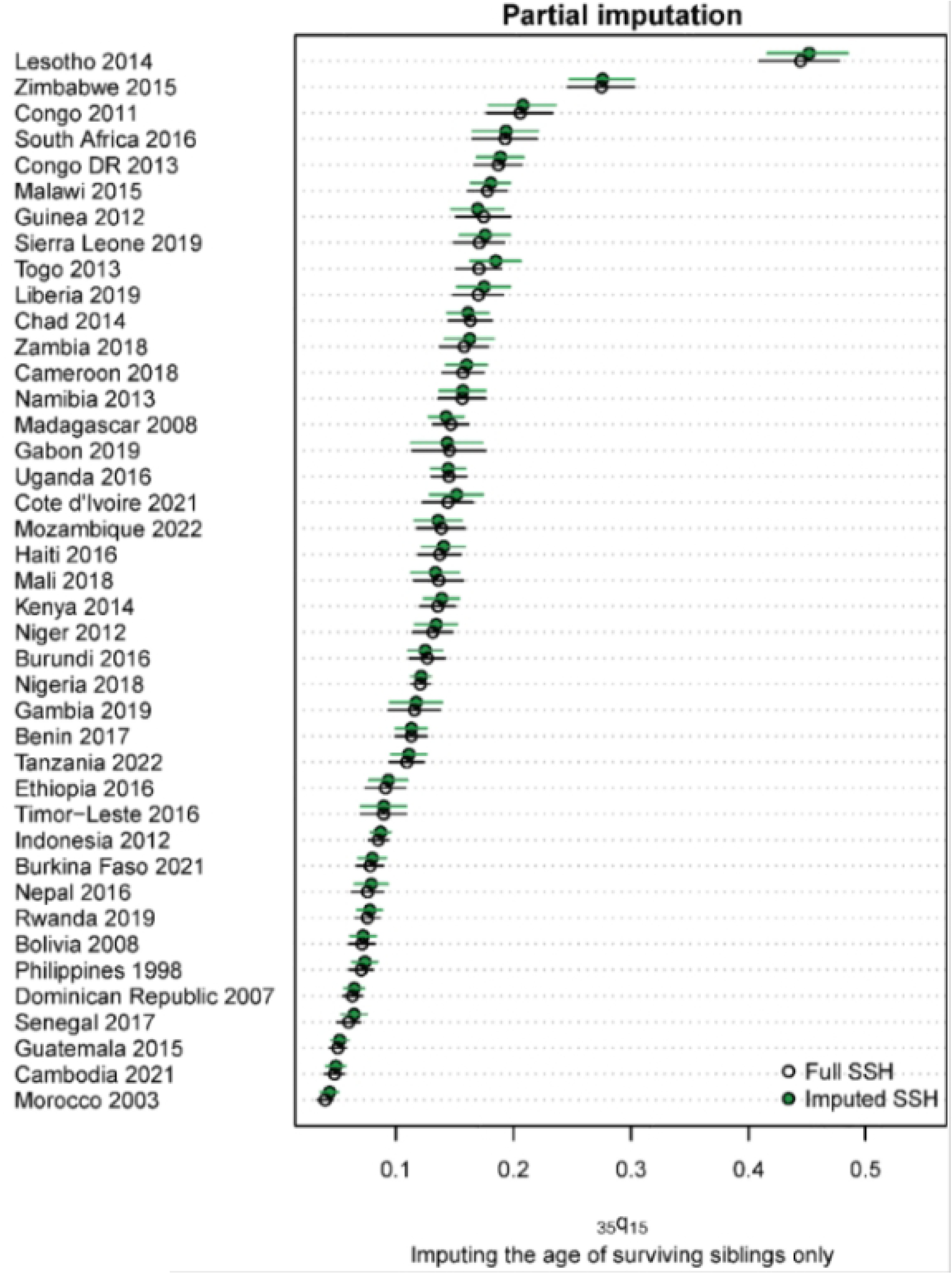
Mean number of siblings ever born, tabulated by cohort of respondents, last two DHS surveys with SSH.

## A.2. Evaluation of the quality of age reporting in the MPS

We used the attraction to round digits when respondents reported their own age as a first indicator of data quality in the MPS. Table S2 presents two indices: the Whipple’s index, assessing the preference for ages ending with the digits 0 and 5, and the Myers’ composite index, capturing heaping on and aversion for all digits from 0 to 9 (45,46). These indices were computed for the MPS and compared to values extracted from previous census DHS or MICS.

In the three countries, heaping on round digits is more pronounced in the MPS than in the face-to-face surveys, with the exception of the EHCVM study arm in Burkina Faso. For example, in Malawi, the Myers index value for the MPS [20] is double that of the face-to-face survey [9.9]. This is consistent with observations made for MPS conducted in other African countries (Côte d’Ivoire, Ghana, Rwanda, Senegal) (21).

The quality of age reporting varies between the two study arms in Burkina Faso and the two provinces in DRC. In Burkina Faso, the Myers index for the EHCVM arm is 18%, against 33% in the RDD arm. The Whipple index also suggests that age misreporting errors are more frequent in the RDD arm. In DRC, both the Myers and Whipple indicators highlight significantly higher accuracy in age reporting in Kinshasa, compared to Nord-Kivu, and this reflects a pattern observed in the face-to-face survey.

**Table S2:**
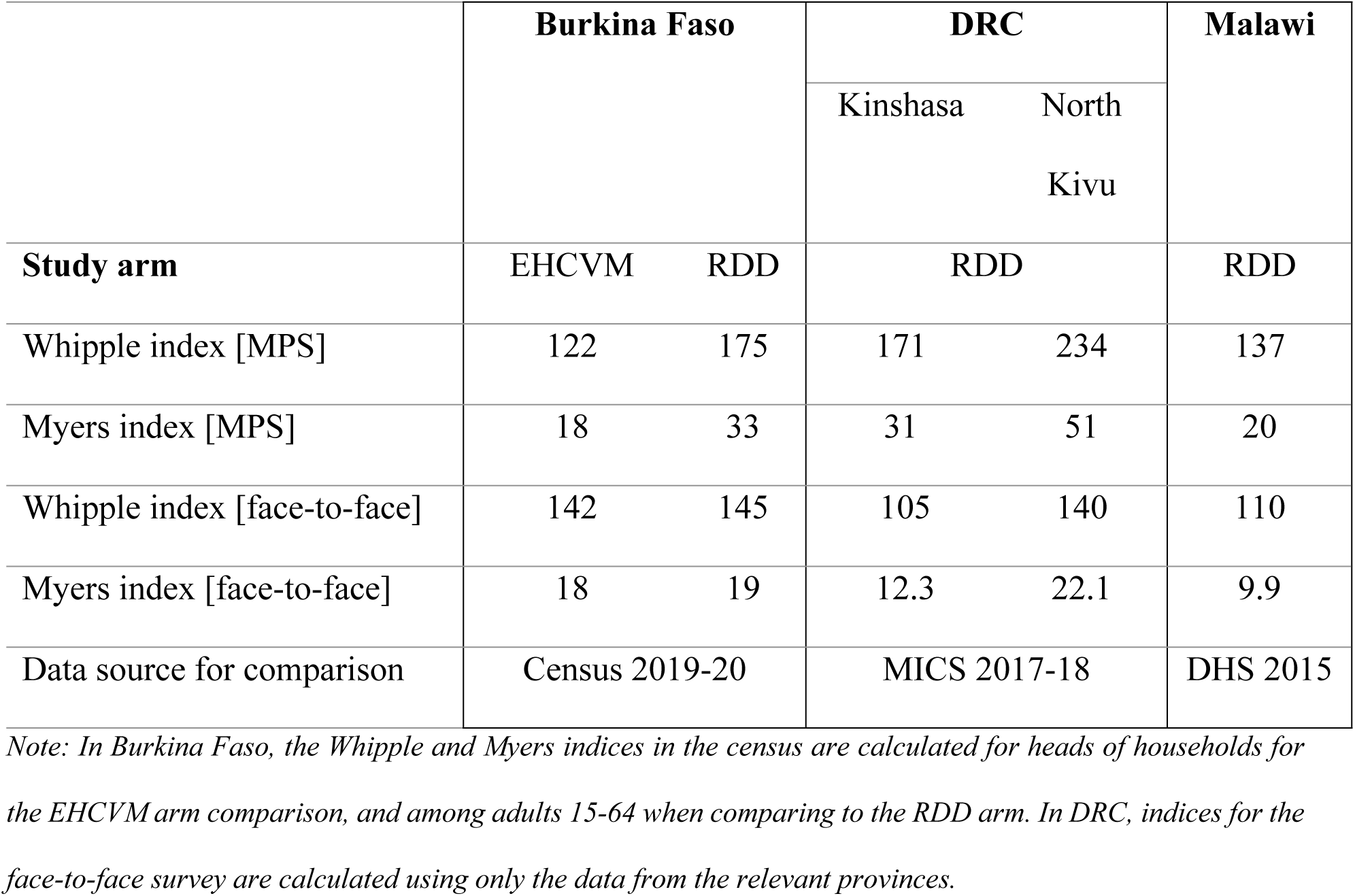
Indices of age heaping in MPS and previous face-to-face surveys, by country.

|  | **Burkina Faso** |  | **DRC** |  | **Malawi** |
| --- | --- | --- | --- | --- | --- |
|  |  |  | Kinshasa | North Kivu |  |
| **Study arm** | EHCVM | RDD | RDD |  | RDD |
| Whipple index [MPS] | 122 | 175 | 171 | 234 | 137 |
| Myers index [MPS] | 18 | 33 | 31 | 51 | 20 |
| Whipple index [face-to-face] | 142 | 145 | 105 | 140 | 110 |
| Myers index [face-to-face] | 18 | 19 | 12.3 | 22.1 | 9.9 |
| Data source for comparison | Census 2019-20 |  | MICS 2017-18 |  | DHS 2015 |

*Note: In Burkina Faso, the Whipple and Myers indices in the census are calculated for heads of households for the EHCVM arm comparison, and among adults 15-64 when comparing to the RDD arm. In DRC, indices for the face-to-face survey are calculated using only the data from the relevant provinces.*

## A.3. Estimating sub-national mortality rates for the provinces of Kinshasa and North-Kivu

Because the adult mortality rates from the WPP refer to the national level, while the sibling-based estimates in DRC refer to two provinces only, we re-scaled the WPP estimates to obtain a reference for Kinshasa and North Kivu. We proceeded as follows:

i. we used the Brass logit system [which has two parameters α – capturing the level of mortality – and β – capturing the age pattern] and assumed that β equals to 1;
ii. we calculated under-five mortality for Kinshasa and North Kivu from the latest DHS conducted in DRC, and computed *Y[5]*, as the logit of survivorship, as follows: Y[5] = -0.5 ln[l_5_/[1-l_5_].
iii. we calculated the logits of the l_x_ values in the reference [national] life table from the WPP.
iv. We estimated the adult mortality levels in Kinshasa and North Kivu, using the formula below;

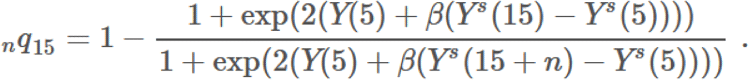

where β is set at 1, *Y[5]* refers to the logits of the DHS-based survivorship in childhood and *Y^s^[5], Y^s^[15]* and *Y^s^[15 + n]* refer to the logits of the survivorship for various age groups in the national WPP life table serving as our reference. This amounts to assuming that the age pattern of mortality in the WPP life table also applies to the two provinces, but the level of mortality is defined by the province-specific estimates of under-five mortality in the DHS.

## A.4 Additional tables

**Table S3.** Questions used to collect summary sibling histories in MPS.

|  |  |  |  |
| --- | --- | --- | --- |
| SSH1 | Now I would like to ask you some questions about your brothers and sisters who are born to your biological mother.<br>Can you please tell me how many sisters were born to your biological mother? This includes sisters who are living with you, sisters who not living with you and those who have died?<br>INSTRUCTION: assist the respondent with a suggestion to enumerate sisters by listing their first names | 0<br>NUMBER .....<br>DON'T KNOW | SSH6 |
| SSH2 | How many of these sisters are no longer with us today [have passed away]? | 0<br>NUMBER<br>DK | SSH6 |
| SSH3 | How many of your sisters have passed away since the beginning of 2019? | 0<br>NUMBER<br>DK | SSH6 |
| SSH4 | Can you give me the first name for these sisters? | S1 FName:<br>S2 FName:<br>S3 FName: |  |
| SSH5 | In which year/month did [NAME] die? | S1 Year:<br>S1 Month: | D<br>LOOP |
|  |  | S2 Year<br>S2 Month<br>S3 Year:<br>S3 Month: | D<br>LOOP<br>D<br>LOOP |
| SSH6 | Now I would like to ask you some questions about your brothers who are born to your biological mother.<br>Can you please tell me how many brothers you have or ever had? Please include brothers who are living with you, sisters who not living with you, and those who have died.<br>INSTRUCTION: assist the respondent with a suggestion to enumerate brothers by listing their first names | 0<br>NUMBER<br>DK |  |
| SSH7 | How many of these brothers are no longer with us today [have passed away]? | 0<br>NUMBER<br>DK |  |
| SSH8 | How many of your brothers have passed away since the beginning of 2019? | 0<br>NUMBER: |  |
| SSH9 | Can you give me the first name for those brothers? | B1 FName:<br>B2 FName:<br>B3 FName: |  |
| SSH10 | In which year/month did [NAME] die? | B1 Year:<br>B1 Month: | D<br>LOOP |
|  |  | B2 Year |  |
|  |  | B2 Month | D<br>LOOP |
|  |  | B3 Year: |  |
|  |  | B3 Month: | D<br>LOOP |

Note: In DRC, we did not collect information on ages at death. In Malawi, we did not collect information disaggregated by sex.

**Table S4:** Information on siblings collected in each country in the MPS.

|  |  | Burkina Faso | DRC | Malawi |
| --- | --- | --- | --- | --- |
| SSH1 | Total number of siblings | ✓ | ✓ | ✓ |
| SSH1_sex | Total number of siblings<br>disaggregated by sex | ✓ | ✓ |  |
| SSH2 | Total number of deceased<br>siblings | ✓ | ✓ | ✓ |
| SSH2_sex | Total number of deceased<br>siblings disaggregated by sex | ✓ | ✓ |  |
| SSH3 | Total number of siblings who<br>have died since the<br>beginning of 2019 | ✓ | ✓ | ✓ |
| SSH3_sex | Sex of siblings who have<br>died since the beginning of<br>2019 | ✓ | ✓ | ✓ |
| SSH5_year | Year of death [for deaths in or<br>after 2019] | ✓ | ✓ | ✓ |
| SSH5 month | Month of death [for deaths in or after 2019] | ✓ | ✓ | ✓ |
| SSH5 age at death | Age at death [for deaths in or after 2019] | ✓ |  | ✓ |

**Table S5:**
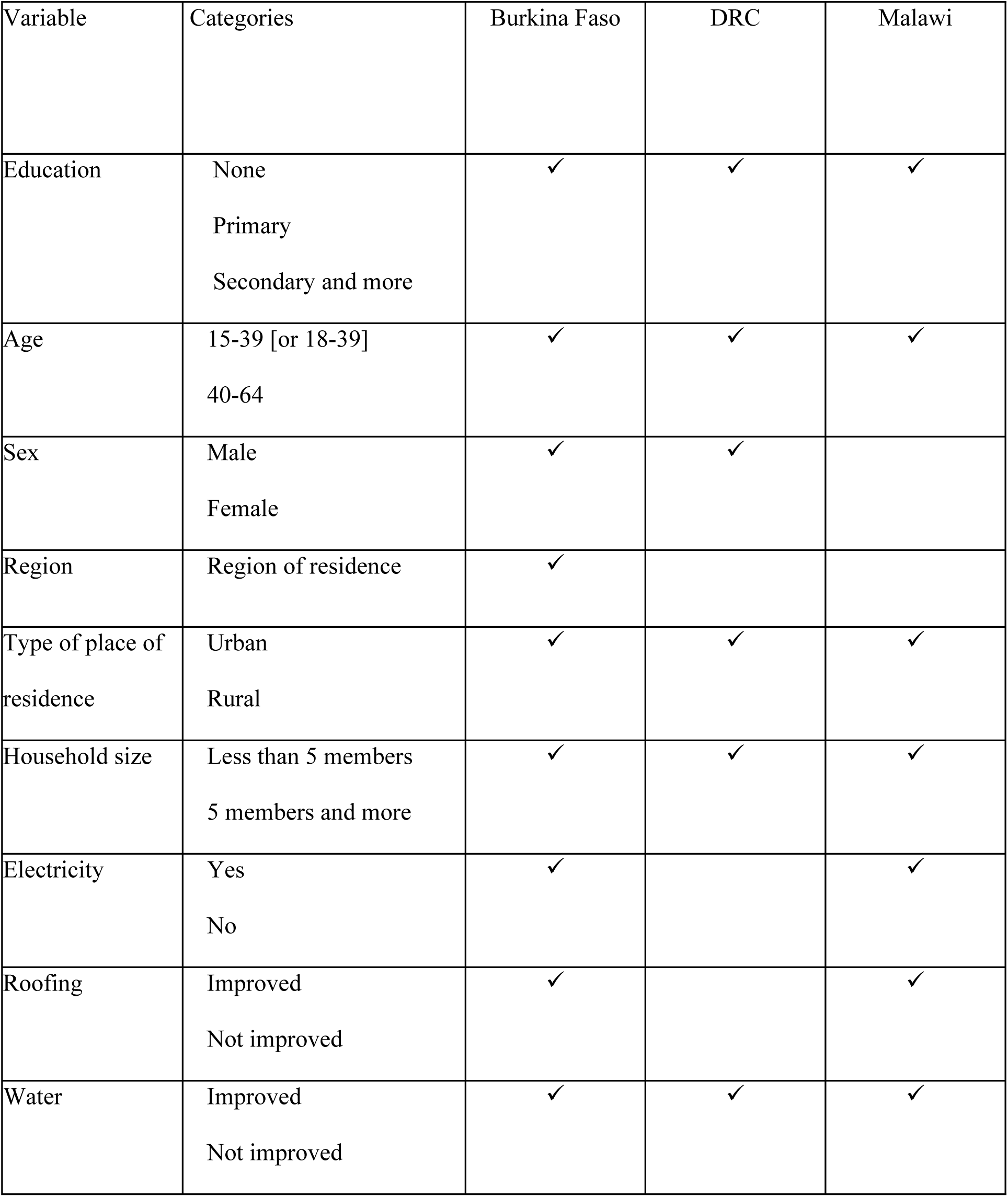
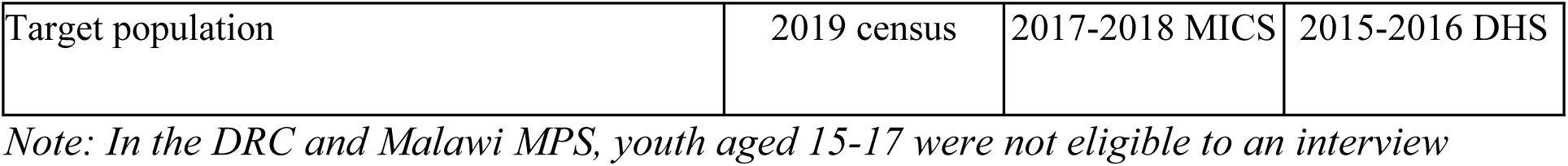
Information collected in MPS and used for calculating weights in each country.

| Variable | Categories | Burkina Faso | DRC | Malawi |
| --- | --- | --- | --- | --- |
| Education | None | ✓ | ✓ | ✓ |
|  | Primary |  |  |  |
|  | Secondary and more |  |  |  |
| Age | 15-39 [or 18-39] | ✓ | ✓ | ✓ |
|  | 40-64 |  |  |  |
| Sex | Male | ✓ | ✓ |  |
|  | Female |  |  |  |
| Region | Region of residence | ✓ |  |  |
| Type of place of residence | Urban | ✓ | ✓ | ✓ |
|  | Rural |  |  |  |
| Household size | Less than 5 members | ✓ | ✓ | ✓ |
|  | 5 members and more |  |  |  |
| Electricity | Yes | ✓ |  | ✓ |
|  | No |  |  |  |
| Roofing | Improved | ✓ |  | ✓ |
|  | Not improved |  |  |  |
| Water | Improved | ✓ | ✓ | ✓ |
|  | Not improved |  |  |  |

|  |  |  |  |
| --- | --- | --- | --- |
| Target population | 2019 census | 2017-2018 MICS | 2015-2016 DHS |

*Note: In the DRC and Malawi MPS, youth aged 15-17 were not eligible to an interview*

**Table S6:** Composition of the MPS sample in Burkina Faso, by study arm, compared with the population enumerated in the 2019-2020 census.

|  | **EHCVM arm [heads of 2019-20** |  |  |  | **RDD arm [population aged 15- 2019-20** |  |  |  |
| --- | --- | --- | --- | --- | --- | --- | --- | --- |
|  | **households]** |  |  |  | **64 years old]** |  |  |  |
| **Respondents** | Unweighted | Weighted |  |  | Unweighted | Weighted |  |  |
| **characteristics** | % | % | 95% CI | % | % | % | 95% CI | % |
| ***Sex*** |  |  |  |  |  |  |  |  |
| Men | 76.7 | 84.0 | [82.0-85.7] | 84.0 | 34.3 | 46.2 | [44.7-47.8] | 46.3 |
| Women | 23.3 | 16.0 | [14.3-18.0] | 16.0 | 65.7 | 53.8 | [52.2-55.3] | 53.7 |
| ***Age group*** |  |  |  |  |  |  |  |  |
| 15-29 | 17.5 | 22.9 | [20.7-25.1] | 22.6 | 41.5 | 43.7 | [42.2-45.3] | 43.7 |
| 30-49 | 52.0 | 49.3 | [46.8-51.9] | 49.1 | 46.8 | 42.2 | [40.7-43.8] | 42.2 |
| 50 &+ | 30.6 | 27.8 | [25.6-30.2] | 28.3 | 11.7 | 14.1 | [13.0-15.2] | 14.1 |
| ***Education*** |  |  |  |  |  |  |  |  |
| None | 52.0 | 69.5 | [67.1-71.8] | 72.2 | 44.9 | 67.6 | [66.1-69.0] | 69.8 |
| Primary | 20.2 | 11.2 | [9.7-12.9] | 10.0 | 16.5 | 11.5 | [10.5-12.5] | 9.6 |
| Secondary | 21.4 | 13.7 | [12.1-15.6] | 13.5 | 29.5 | 15.7 | [14.6-16.8] | 15.9 |
| Tertiary and higher | 6.4 | 5.6 | [4.5-6.9] | 4.2 | 9.0 | 5.2 | [4.6-5.9] | 3.6 |
| ***Marital status*** |  |  |  |  |  |  |  |  |
| Married | 83.6 | 83.3 | [81.3-85.1] | 84.2 | 75.4 | 77.5 | [76.2-78.8] | 72.5 |
| Widowed | 5.5 | 5.0 | [4.0-6.2] | 1.1 | 3.3 | 3.6 | [3.1-4.3] | 3.4 |
| Divorced/sep | 0.8 | 0.7 | [0.4-1.3] | 5.6 | 0.8 | 0.7 | [0.5-1.0] | 0.9 |
| Single | 10.1 | 11.0 | [9.5-12.7] | 9.1 | 20.4 | 18.2 | [17.0-19.4] | 23.2 |
| ***Type of place of residence*** |  |  |  |  |  |  |  |  |
| Ouagadougou | 11.1 | 13.0 | [11.4-14.8] | 14.4 | 15.8 | 11.5 | [10.6-12.4] | 14.5 |
| Bobo-Dioulasso | 6.1 | 3.4 | [2.6-4.4] | 5.0 | 6.7 | 3.8 | [3.3-4.4] | 5.2 |
| Other towns | 40.2 | 13.5 | [12.0-15.3] | 11.9 | 14.3 | 9.3 | [8.5-10.2] | 11.4 |
| Rural areas | 42.6 | 70.1 | [67.7-72.3] | 68.7 | 63.2 | 75.4 | [74.1-76.6] | 69.0 |
| ***Region of residence*** |  |  |  |  |  |  |  |  |
| Boucle du Mouhoun | 7.1 | 10.5 | [9.0-12.2] | 9.6 | 6.8 | 10.6 | [9.6-11.7] | 9.5 |
| Cascades | 7.4 | 4.3 | [3.4-5.5] | 3.9 | 4.1 | 4.1 | [3.5-4.8] | 4.1 |
| Centre | 15.7 | 16.8 | [14.9-18.8] | 18.4 | 24.3 | 13.4 | [12.5-14.5] | 18.0 |
| Centre-Est | 7.8 | 7.9 | [6.6-9.4] | 7.7 | 6.9 | 8.1 | [7.3-9.1] | 7.4 |
| Centre-Nord | 7.9 | 7.6 | [6.4-9.1] | 7.0 | 9.2 | 8.5 | [7.6-9.4] | 7.3 |
| Centre-Ouest | 6.8 | 7.4 | [6.1-8.9] | 7.8 | 7.6 | 8.3 | [7.5-9.2] | 8.0 |
| Centre-Sud | 5.6 | 4.5 | [3.6-5.7] | 4.0 | 4.3 | 4.6 | [3.9-5.3] | 3.9 |
| Est | 7.1 | 8.5 | [7.2-10.1] | 7.5 | 4.0 | 8.3 | [7.5-9.3] | 7.8 |
| Hauts-Bassins | 12.7 | 12.0 | [10.4-13.7] | 11.8 | 14.2 | 11.0 | [10.1-11.9] | 12.0 |
| Nord | 6.5 | 7.7 | [6.4-9.2] | 7.8 | 7.4 | 8.5 | [7.6-9.4] | 8.1 |
| Plateau-Central | 6.3 | 4.5 | [3.6-5.7] | 4.6 | 6.2 | 4.8 | [4.2-5.6] | 4.7 |
| Sahel | 2.9 | 3.8 | [2.9-4.9] | 5.3 | 1.7 | 5.4 | [4.7-6.2] | 4.6 |
| Sud-Ouest | 6.2 | 4.4 | [3.5-5.6] | 4.8 | 3.4 | 4.3 | [3.7-5.0] | 4.6 |
| ***Household size*** |  |  |  |  |  |  |  |  |
| 1-4 members | 21.5 | 32.9 | [30.5-35.3] | 49.4 | 28.5 | 39.2 | [37.6-40.7] | 49.4 |
| 5-8 members | 43.0 | 37.4 | [34.9-39.9] | 36.5 | 41.9 | 33.7 | [32.3-35.3] | 36.5 |
| 9+ | 35.4 | 29.7 | [27.4-32.1] | 14.1 | 29.6 | 27.1 | [25.7-28.6] | 14.1 |

Note: The percentages in the columns referring to the census are specific to each arm because the EHCVM study arm is compared to the population of heads of households in the census, while the RDD sample is compared to the population aged 15-64.

**Table S7:**
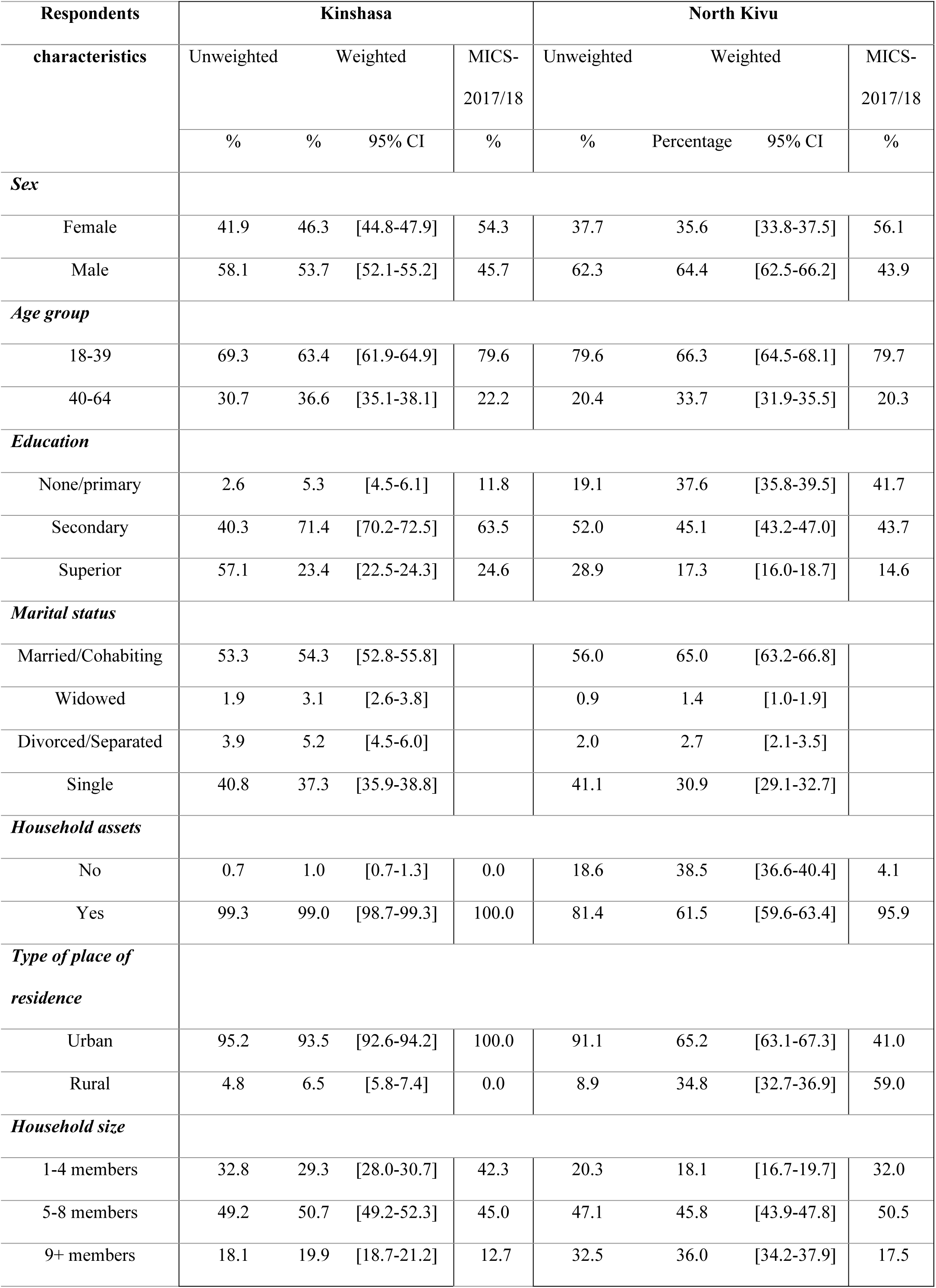
Composition of the MPS sample in DRC, by province, compared with the population enumerated in the MICS-2017-2018 survey.

**Table S8:** Composition of the MPS sample in Malawi, compared with the population enumerated in the 2018 census.

| Respondents characteristics | Unweighted | Weighted |  | 2018 |
| --- | --- | --- | --- | --- |
|  | % | % | 95% CI | census<br>% |
| ***Sex*** |  |  |  |  |
| Female | 47.5 | 46.9 | [45.6-48.2] | 46.9 |
| Male | 52.5 | 53.1 | [51.8-54.4] | 53.2 |
| ***Age group*** |  |  |  |  |
| 18-49 | 92.3 | 85.1 | [84.1-86.1] | 77.3 |
| 50+ | 7.7 | 14.9 | [13.9-15.9] | 22.7 |
| ***Education*** |  |  |  |  |
| None/primary | 22.5 | 44.3 | [43.0-45.6] | 72.7 |
| Secondary | 50.9 | 49.1 | [47.8-50.4] | 23.4 |
| Superior | 26.6 | 6.6 | [5.9-7.3] | 3.9 |
| ***Marital status*** |  |  |  |  |
| Married/Cohabiting | 58.0 | 64.8 | [63.6-66.0] | 66.1 |
| Widowed | 2.4 | 3.5 | [3.0-4.0] | 7.2 |
| Divorced/Separated | 7.2 | 8.8 | [8.1-9.6] | 7.9 |
| Single | 32.4 | 22.9 | [21.8-23.9] | 18.8 |
| ***Type of place of residence*** |  |  |  |  |
| Urban | 26.0 | 18.0 | [17.1-19.0] | 16.9 |
| Rural | 74.0 | 82.0 | [81.0-82.9] | 83.1 |
| ***Region*** |  |  |  |  |
| Central | 39.9 | 40.7 | [39.4-42.0] | 43.1 |
| Northern | 15.5 | 12.9 | [12.1-13.9] | 12.2 |
| Southern | 44.6 | 46.4 | [45.1-47.7] | 44.7 |
| **Household size** |  |  |  |  |
| 1-4 members | 39.3 | 34.4 | [33.2-35.7] | 92.0 |
| 5-8 members | 51.8 | 56.4 | [55.1-57.7] | 7.8 |
| 9+ members | 8.9 | 9.2 | [8.5-9.9] | 0.2 |

Note: The sample is compared to the population aged 18-64 years old in Malawi

**Table S9:**
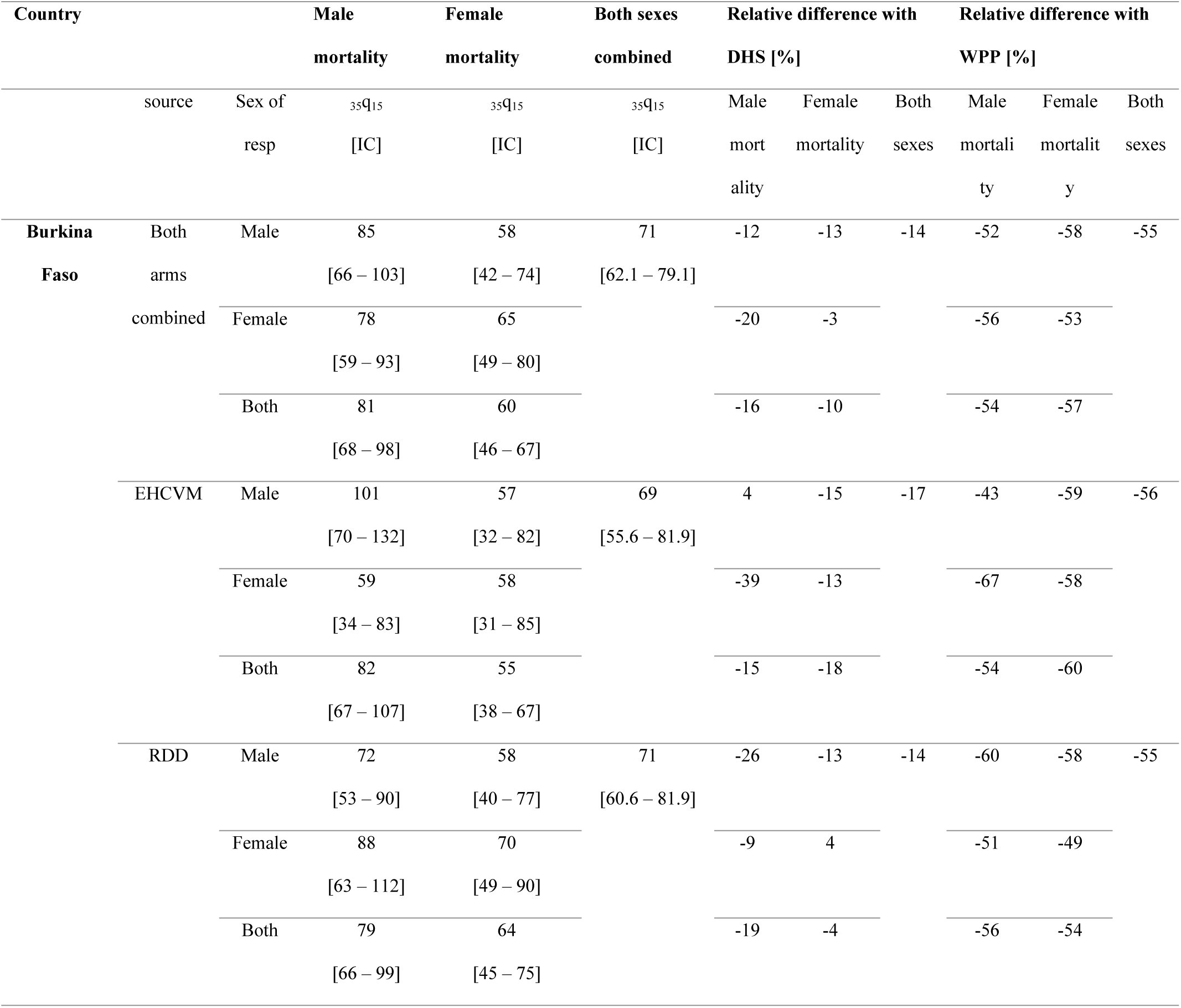
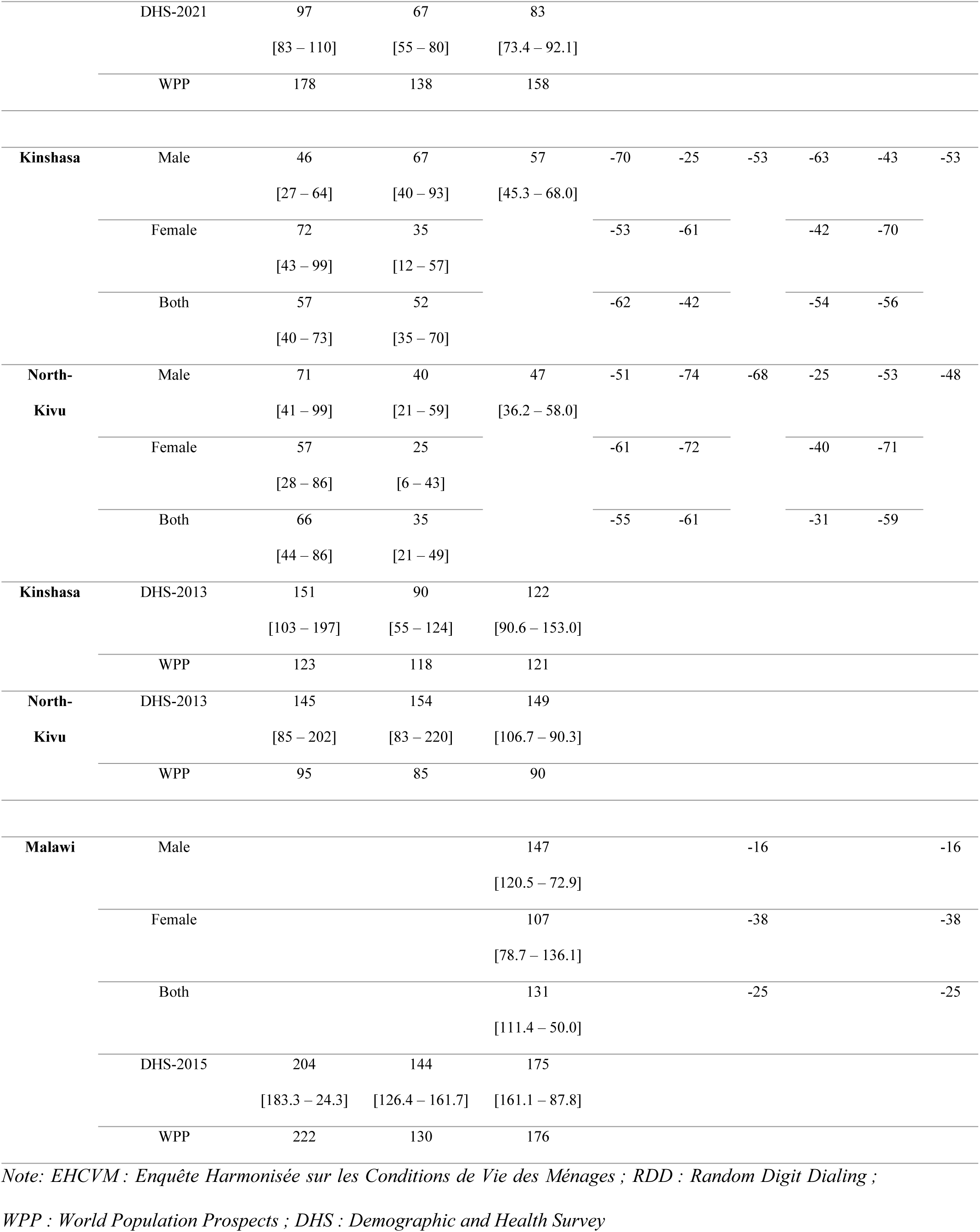
Estimates of the probability _35_q_15_ over the period 0-3 years before data collection, by source and sex of respondent and 95% confidence intervals per country [‱], based on partial imputation.

**Table S10:**
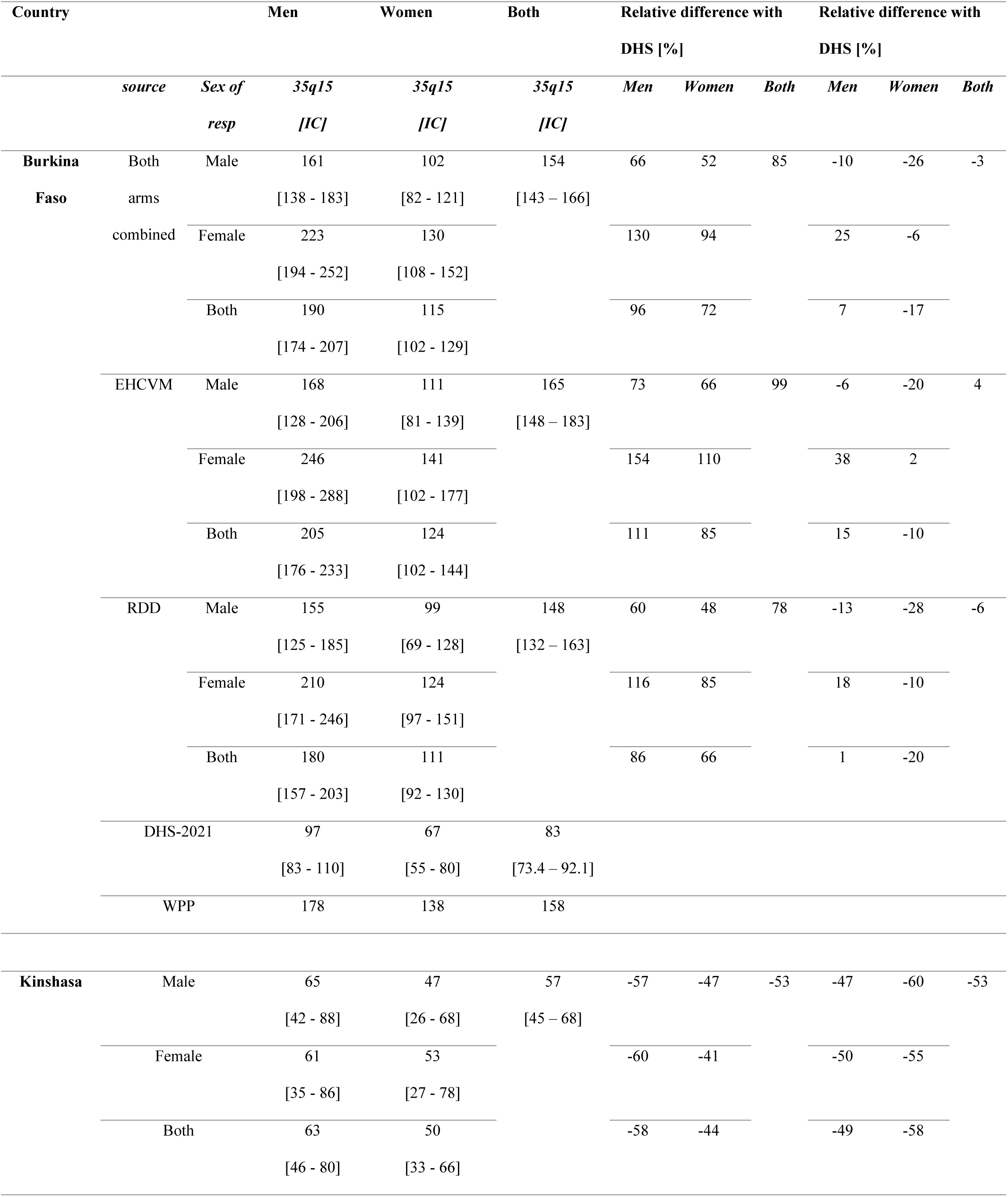
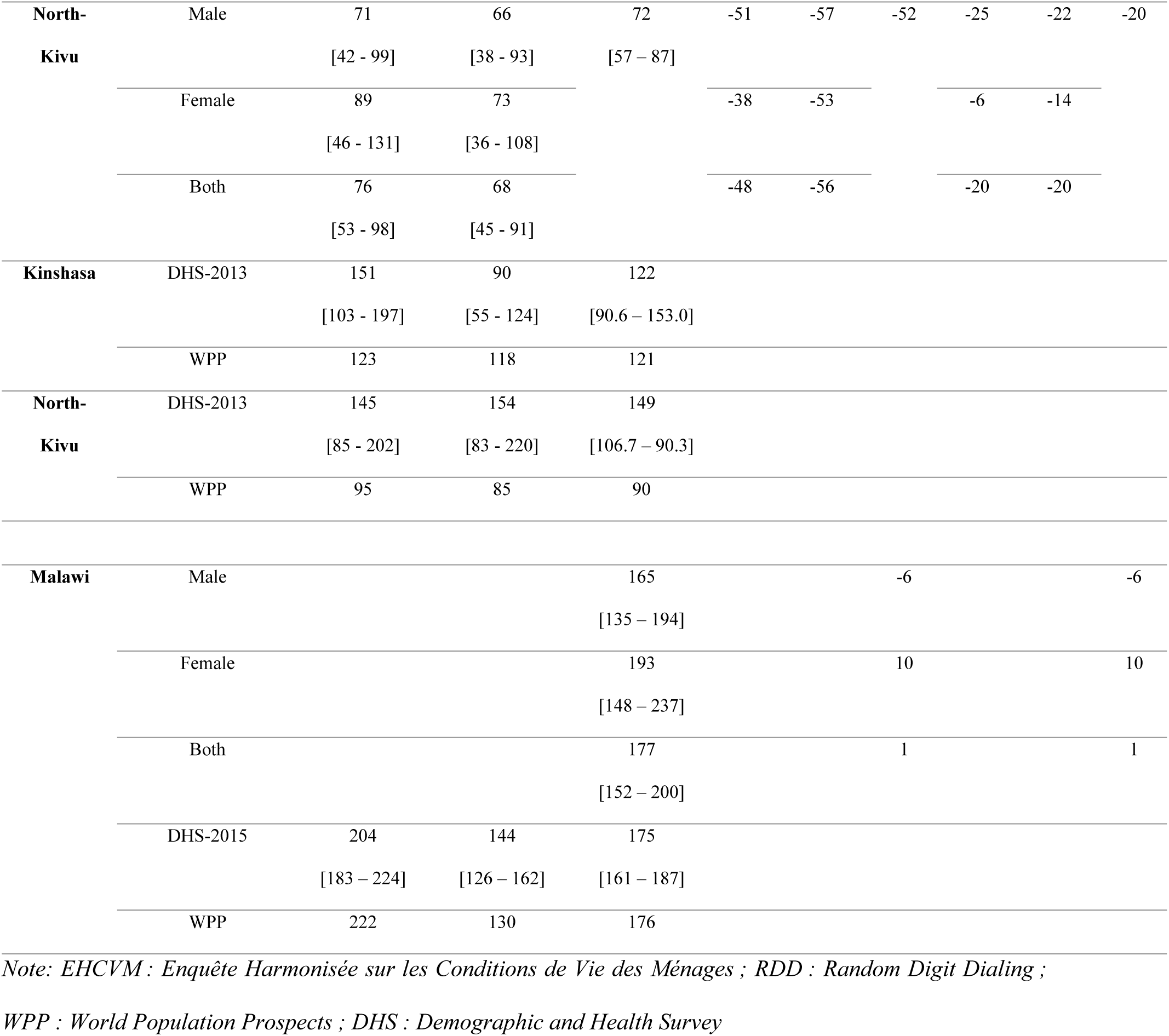
Estimates of the probability _35_q_15_ over the period 0-3 years before data collection, by source and sex of respondent and 95% confidence intervals per country (deaths before age 50 for 1000 adolescents aged 15), based on full imputation.

## 6.2 Additional figures

**Fig S4.**
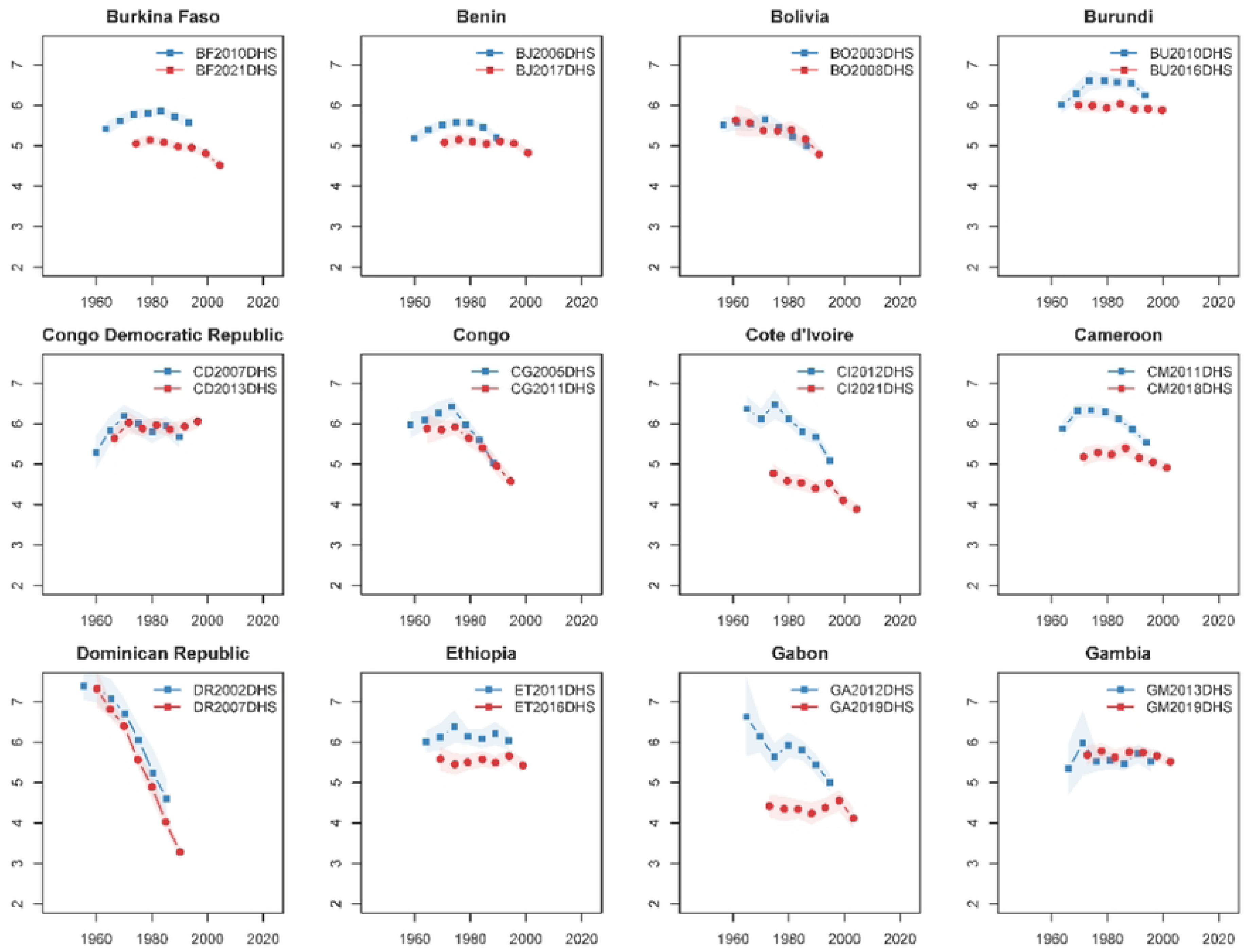
Proportions of surviving siblings at the time of the survey, by age group of respondent and sex of siblings in MPS and DHS, in Burkina Faso and DRC.

**Fig S5.**
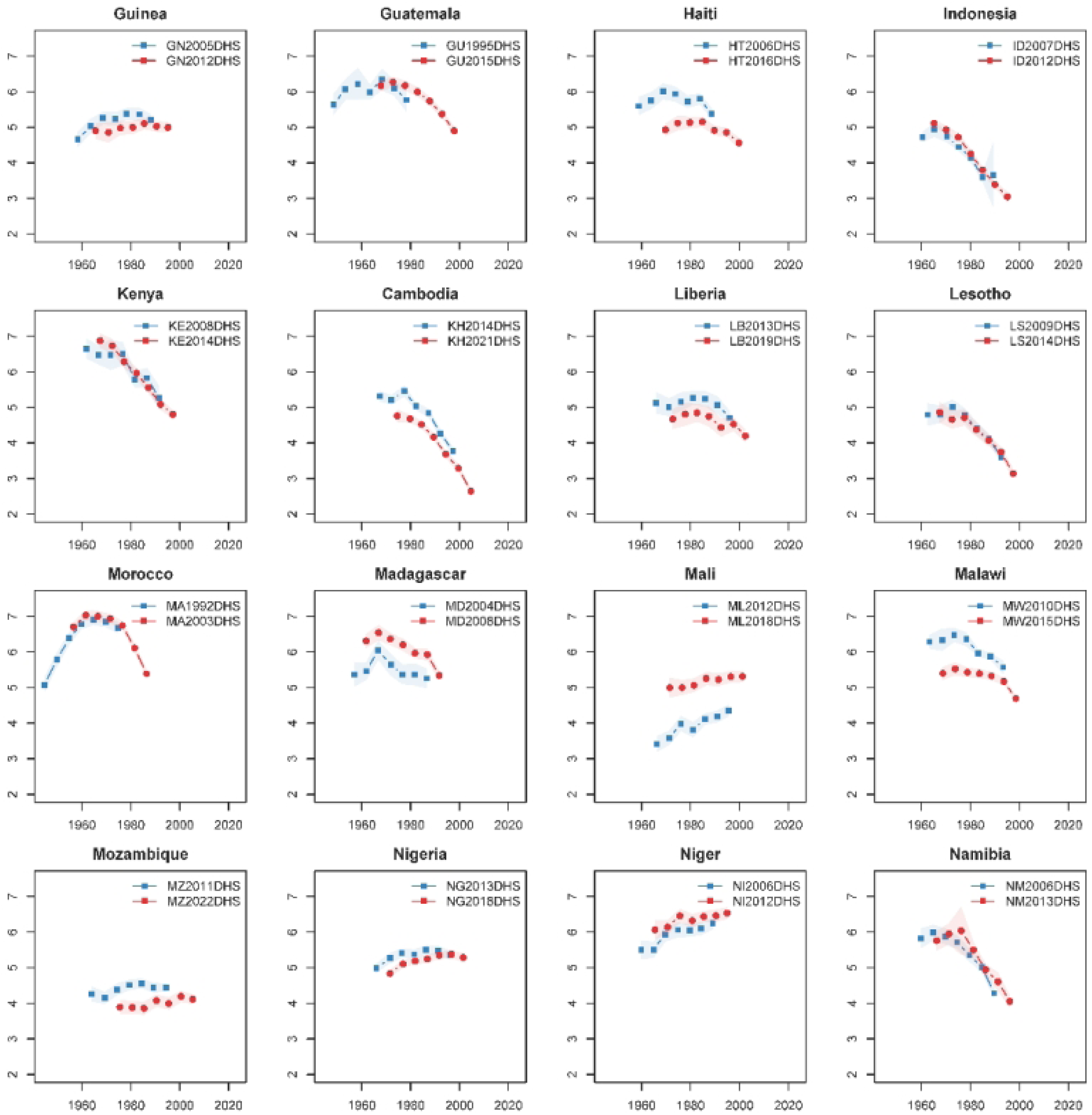
Proportions of recent deaths among all deceased siblings by age group of respondent and sex of sibling [last 3 years in DHS and since January 2019 in MPS] in Burkina Faso and DRC.

**Fig S6:**
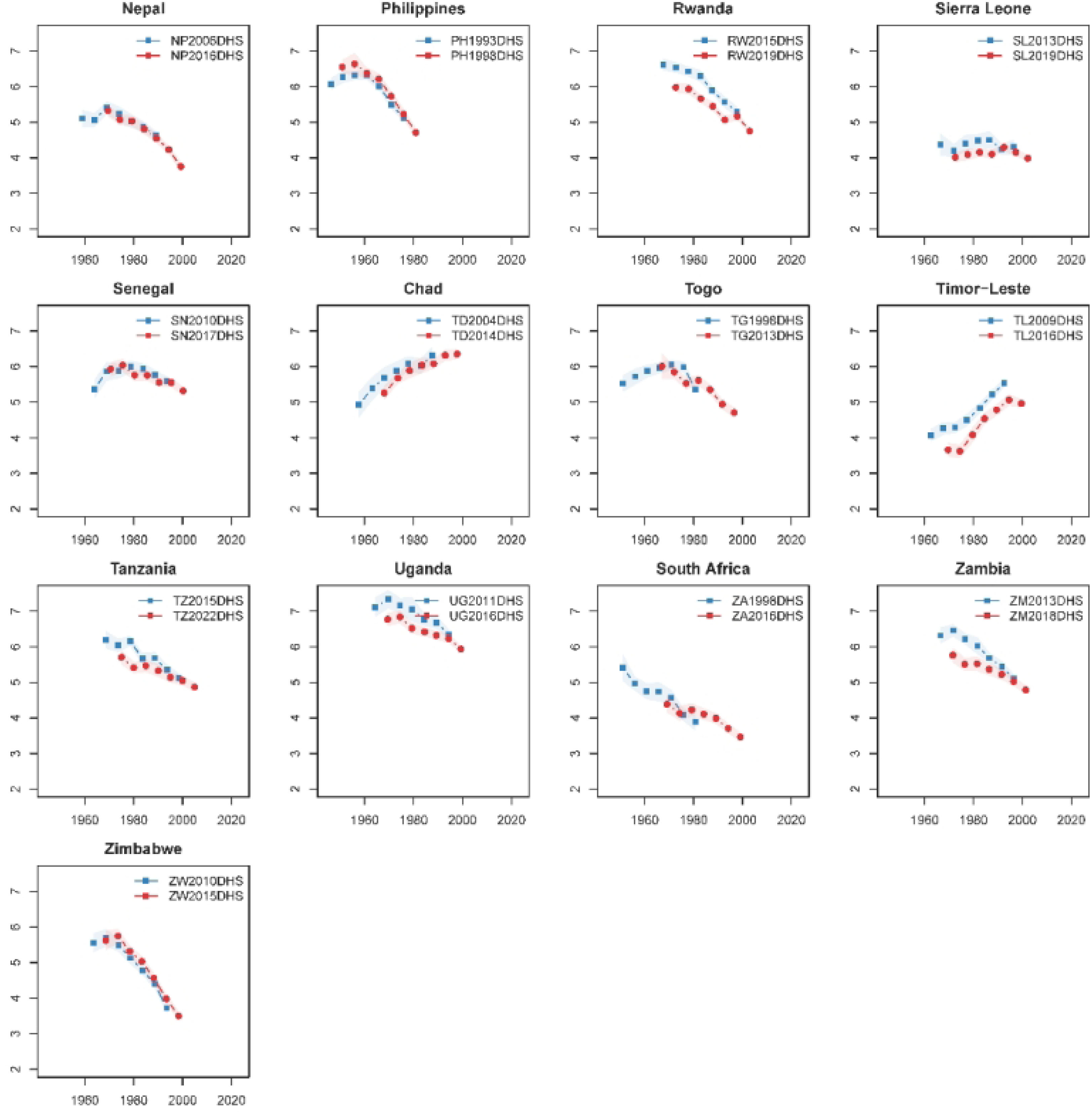
Trends in adult mortality [_35_q_15_] according to SSH in the MPS survey [using partial imputation] or DHS surveys and in the World Population Prospects.

